# Methods of adjustment for non-vaccine interventions in post-licensure vaccine studies in children in sub–Saharan Africa: a systematic review

**DOI:** 10.64898/2026.04.01.26349767

**Authors:** Latif Ndeketa, Natasha Marcella Vaselli, Virginia E. Pitzer, Peter J. Dodd, Daniel Hungerford, Neil French

## Abstract

**Background:** Post-licensure vaccine effectiveness and impact studies provide evidence on how vaccines perform under routine programme conditions in the real world. In sub-Saharan Africa (SSA), vaccine introductions frequently coincide with concurrent public health and social measures that may influence disease risk and transmission. Failure to account for these concurrent interventions may affect the interpretation of vaccine effects.

**Methods:** We conducted a systematic review of post-licensure vaccine effectiveness and impact studies conducted in children under five years of age in SSA. Electronic databases were searched for peer-reviewed studies published between January 2000 and December 2019. Eligible studies used observational designs to estimate vaccine effectiveness or population-level impact. Two reviewers independently screened studies, extracted data, and assessed methodological quality using Joanna Briggs Institute tools. We examined study designs, vaccines evaluated, outcomes assessed, and whether public health and social measures (PHSMs) were measured or adjusted for. A narrative synthesis was undertaken. In addition, we conducted a meta-analysis for rotavirus and pneumococcal conjugate vaccines where we explored the heterogeneity in individual-level effectiveness estimates where designs and outcomes were comparable.

**Results:** Sixty-four studies met the inclusion criteria, covering eight vaccine-preventable diseases. Rotavirus vaccines were most frequently evaluated, followed by pneumococcal conjugate vaccines. Case-control and ecological designs were most common, while cohort and time-series analyses were less frequently used. None of the included studies collected, reported, or adjusted for PHSMs such as nutrition, WASH, or access to healthcare. The implications of this omission varied by pathogen. Rotavirus vaccine effectiveness estimates from comparable individual-level designs were consistent across settings, with no evidence of between-study heterogeneity. Pneumococcal vaccine effectiveness estimates showed substantial heterogeneity, which appeared to reflect differences in outcome definitions, host risk profiles, and study context. Estimates for other vaccines were generally protective in direction, although the magnitude and precision varied across studies.

**Conclusions:** Post-licensure vaccine effectiveness and impact studies in SSA rarely account for concurrent PHSMs. The consequences of this omission are not uniform across vaccines. For some pathogens, effectiveness estimates appear robust to unmeasured contextual change, while for others they are highly sensitive to outcome choice and setting. Future evaluations should prioritise systematic measurement of key PHSMs and consider study designs that better account for time-varying context. Strengthening routine data systems to capture these factors is essential for generating interpretable evidence to inform immunisation policy.

**Funding:** MRC Discovery Medicine North (DiMeN) Doctoral Training Partnership (UKRI), National Institute for Health and Care Research (NIHR) Global Health Research Group on Gastrointestinal Infections and Wellcome through the core grant to the Malawi-Liverpool-Wellcome Research Programme.

## Introduction

### Background of vaccine preventable diseases

Vaccination remains one of the most effective public health interventions for reducing childhood morbidity and mortality. National immunisation programmes in sub-Saharan Africa (SSA) have progressively introduced vaccines targeting major childhood pathogens including tuberculosis, diphtheria, tetanus, pertussis, poliomyelitis, pneumococcal disease, rotavirus diarrhoea, measles, and more recently typhoid and malaria(1). These vaccines have contributed to substantial declines in vaccine-preventable diseases across the region(2–5). Despite this progress, vaccine-preventable infections continue to account for a substantial burden of child mortality, with the majority of deaths occurring in low-income settings where health system constraints and high transmission intensity persist(4). The introduction of many vaccines has occurred alongside broader improvements in child health, including expanded malaria control, improved nutrition, HIV prevention programmes, and water, sanitation and hygiene (WASH) interventions. These concurrent public health and social measures (PHSMs) may influence disease burden and complicate attribution of observed reductions to vaccination alone when evaluating vaccine effectiveness or impact.

### Overview of post-licensure study designs for vaccine evaluation

Pre-licensure clinical trials evaluate vaccine safety and efficacy under controlled conditions and are widely regarded as the gold standard for establishing direct protective effects(6). Individually randomised trials estimate vaccine efficacy in vaccinated individuals compared with controls, while cluster-randomised trials can also capture indirect protection by allocating communities or geographical areas to vaccination or control groups(6–8). These trials are designed primarily to support licensure and therefore typically include strict eligibility criteria and relatively defined endpoints, which may not fully reflect vaccine performance in routine use.

Post-licensure studies therefore provide essential evidence on how vaccines perform once introduced into national immunisation programmes across diverse populations and epidemiological settings. These studies can quantify direct protection in vaccinated individuals as well as broader population effects arising from reduced transmission, including indirect and overall programme impact(9,10). A range of observational designs are used for this purpose, including ecological analyses, time-series approaches, case-control studies, cohort studies and screening methods. Such designs are often necessary because withholding an approved vaccine from populations in need would raise ethical concerns. Observational evaluations nevertheless introduce additional methodological challenges, including confounding from concurrent PHSMs and broader changes in population health that occur alongside vaccine introduction.

### Challenges in evaluating vaccine effectiveness

There have been observed differences in vaccine effectiveness for particular vaccines in different geographical settings, depending on differing coverage of other interventions for the same disease(22–25). There is no agreed methodological standard for the design of post-licensure vaccine effectiveness and impact studies. The modifying effects of disease related PHSMs could be important when evaluating impact of vaccination programmes. Furthermore, the epidemiology of other diseases, health interventions, health systems, surveillance efforts and societal developments are continually changing and are often difficult to quantify via the routine surveillance systems common in Sub-Saharan Africa, yet these factors may all confound impact measures. This makes it a necessity to use appropriate methods when estimating vaccine-attributable effects, particularly when studying nonspecific endpoints such as mortality and symptomatic health care utilisation. In this review, we examined the principal study designs used in post-licensure vaccine effectiveness and impact studies and assessed the extent to which these studies accounted for concurrent PHSMs.

## Methods

### Design

This systematic review was conducted using the Preferred Reporting Items for Systematic Reviews and Meta-Analyses (PRISMA) guidelines. It aimed to investigate the types of study designs used in post-licensure vaccine evaluations and whether they actively considered PHSMs, and if yes, how they accounted for them in study design and/or adjusted for them in analyses. The review was registered on PROSPERO for systematic reviews (registration number: CRD42023436851)

### Search strategy

A comprehensive search was conducted in electronic databases to identify relevant post-licensure vaccine studies that were published in peer-reviewed journals from the year 2000 to 2019. This was done to exclude the post-licensure studies related to COVID-19 vaccines. The electronic databases included in the search were PubMed Central, EMBASE, MEDLINE and CINAHL. In addition, to identify missing papers and non-academic literature, Google Scholar was searched and citation searching of reference list of included papers was also conducted. Review articles on vaccine impact and effectiveness studies were also used to search the references for relevant publications. The search was restricted to human studies, in English language and excluded pre-prints and review articles. The search dates were from 24/05/2023 to 27/05/2023.

To develop a comprehensive search query we used Boolean operators “AND”, “OR” and “NOT” to combine Medical Subject Headings (MeSH) terms and keywords to develop a final search string: (((“Vaccination”[MeSH Terms] OR “Immunization”[MeSH Terms] OR “immuni*”[Title/Abstract] OR “vaccin*”[Title/Abstract] OR “vaccination program*”[Title/Abstract] OR “immunization program*”[Title/Abstract] OR “immunisation program*”[Title/Abstract]) AND “Africa South of the Sahara”[MeSH Terms] AND (“Infant”[MeSH Terms] OR “child*”[Title/Abstract] OR “preschool”[Title/Abstract]) AND (“Program Evaluation”[MeSH Terms] OR “Effectiveness”[Title/Abstract] OR “Impact”[Title/Abstract])) NOT “Review”[Publication Type]) AND ((humans[Filter]) AND (english[Filter]) AND (allinfant[Filter] OR preschoolchild[Filter]) AND (2000:2019[pdat]))

### Study selection and screening

Studies that were identified using the search strategy were imported into Rayyan AI, a web-based tool for managing systematic reviews(26). Rayyan AI was used to create a database of records and was also instrumental in handling the large number of records from multiple databases and de-duplicating the records.

Two blinded reviewers (LN and MV) carried out the two-stage study selection by (1) screening titles and abstracts and (2) the full text review to select articles that met the inclusion and exclusion criteria. Following the unblinding procedure, discrepancies that arose at both stages of study selection were resolved by discussion, re-examination and consensus. Figure 1 shows the outline of the review.

**Figure 1:**
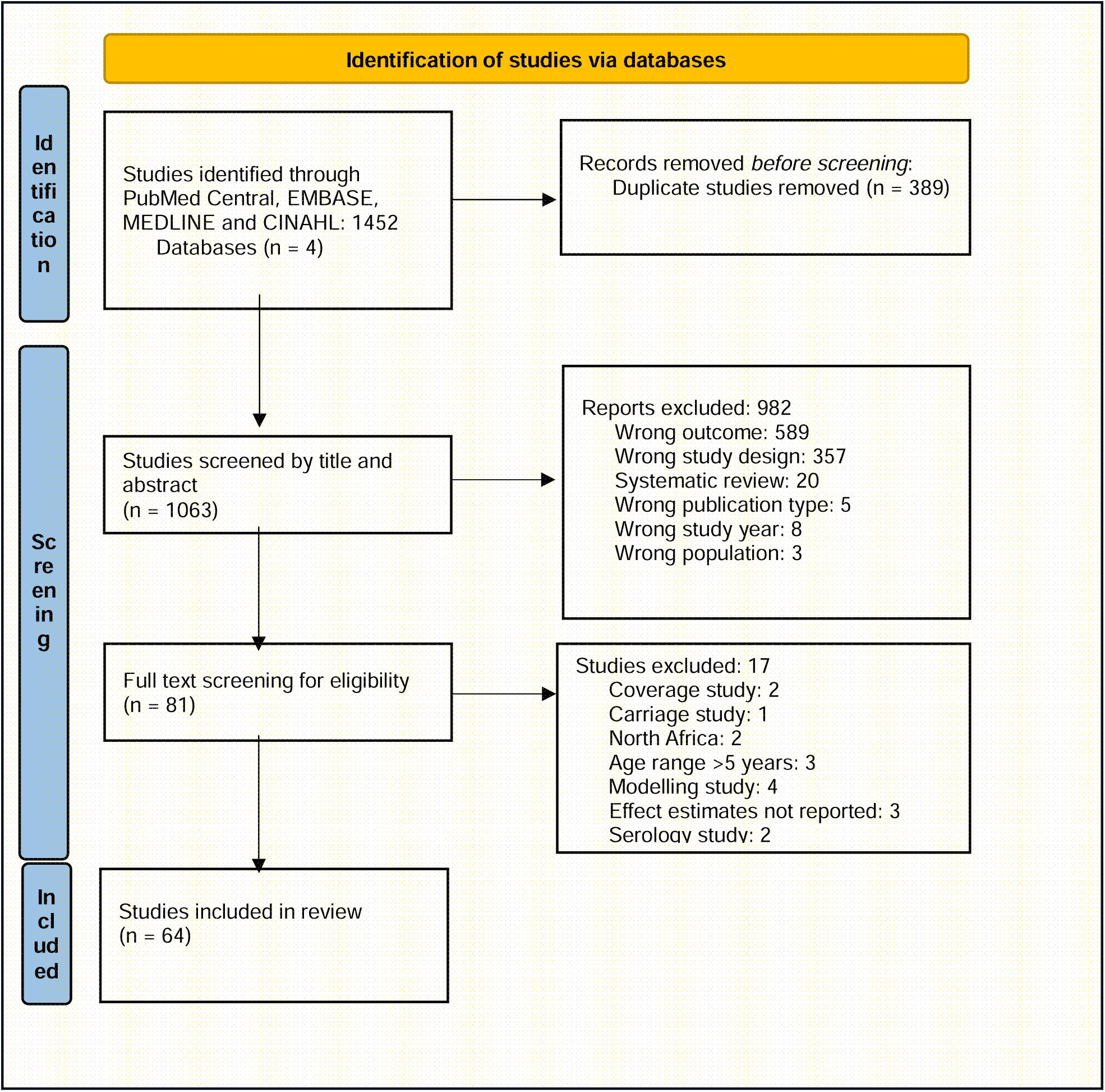
Flow chart selection process (PRISMA)

### Inclusion and exclusion criteria

Studies were included if they were: (1) Articles published in peer reviewed journals for vaccine effectiveness and vaccine impact studies; (2) Used observational methods (e.g. case-control, cohort, time series, ecological and cluster randomized designs); (3) Published between 01 January 2000 and 31 December 2019; (4) Conducted in children living in Africa and (5) were published in English.

Studies were not included if they were: (1) Clinical trials; (2) pre-prints; (3) letters to the editor; (4) conference abstracts; (5) modelling studies that did not use observational data; (6) cost-effectiveness evaluations; (7) systematic reviews; (8) unable to access full articles; (9) serological studies; (10) vaccine coverage studies; (11) carriage studies and (12) pre – post introduction evaluations that used hospital registry data and did not report effect measures.

### Data extraction

A data extraction form was developed using Microsoft Excel to systematically collect relevant data from studies including title, year of publication, journal name, author names, geographical location (country), disease of interest, vaccine of interest, other confounding PHSMs, magnitude of effect of PHSMs, adjustment for PHSMs, adjustment for other vaccines, study design, setting (community/hospital), funder, effect estimates, inclusion criteria, exclusion criteria, primary end points, sample size, study period, follow-up period, study population, data collection methods, herd immunity, follow-up procedures, measures of effect and type of funder.

### Quality assessment of included studies

The Joanna Briggs Institute (JBI) quality assessment tool was used by two independent reviewers (LN and MV) on the studies that met the inclusion criteria. Any conflicts that arose were resolved through re-examination and advice from a third reviewer (DH)

### Data analysis

A qualitative narrative of the reviewed papers was performed to summarise the outcomes given the heterogeneity of the study designs and other outcomes. The characteristics of studies were described in tables based on variables including, year of publication, vaccine of interest, study design, PHSMs assessed and other risk factors. For measures of vaccine effectiveness and impact we report point estimates and range of point estimates across studies. We have included meta-analyses and forest plots of effectiveness in case-control studies by vaccine type to assess between-study heterogeneity. The objective was not to generate pooled vaccine effectiveness estimates, which have been reported in other systematic reviews of specific vaccines. Instead, the analysis was intended to assess how reported effectiveness varies across comparable study designs and settings, and to consider whether differences in analytical approaches and contextual factors may contribute to this variation. Adjusted Odds Ratios and confidence intervals reported from case-control studies were included. We used random effects models with inverse variance method, using the DerSimonian-Laird between study heterogeneity estimator. Heterogeneity was measured using chi-squared (χ2) heterogeneity p-values and I^2^ statistics.

## Results

### Study selection

The search across electronic databases is presented in detail in figure 1 and yielded 64 studies included in the review. The majority were excluded because the study design and outcome did not meet the inclusion criteria.

### JBI quality assessment results

The quality of the methods of included studies was moderate to high across all study designs. Using the Joanna Briggs Institute appraisal tools, none of the 64 studies were classified as poor quality by either reviewer. Most studies clearly defined their study populations, outcomes, confounders, analytical approaches, and applied appropriate statistical methods for their respective designs. However, despite generally strong internal validity within designs, none collected, reported, or adjusted for concurrent PHSMs.

### Study characteristics

Our review included 64 studies evaluating post-licensure vaccine effectiveness across sub-Saharan Africa, revealing patterns in research approaches and focus areas detailed in Table 1.

**Table 1.** Overview of post-licensure study designs for vaccine evaluation.

| Study Design | What it estimates | Strengths | Limitations |
| --- | --- | --- | --- |
| Ecological (including uncontrolled and controlled interrupted time-series)(11–14) | Overall effect or population level impact | Uses routinely collected surveillance data; can capture broad programmatic impact rapidly | Highly vulnerable to confounding from secular trends and concurrent interventions; no individual-level inference |
| Screening method(15) | Overall vaccine effectiveness (VE) using coverage and case vaccination status | Low cost; relatively quick; minimal data requirements | Dependent on accurate coverage and case ascertainment data; prone to misclassification bias; limited to VE estimation |
| Case-control (including test-negative and matched designs) (15,16) | Direct VE estimation at individual level | Efficient for rare outcomes; suitable during outbreaks and immediately after new vaccine introductions; requires smaller sample sizes | Susceptible to selection and recall bias; careful adjustment for confounding needed. Challenging if vaccine is highly effective, due to too few cases and / or if vaccine uptake is very high or low, best implemented when uptake is 20-80%. |
| Cohort studies(17–20) | Direct, indirect, total and overall effect | Strong temporal sequence; can estimate incidence; can assess multiple outcomes | Expensive; follow-up losses; residual confounding by health-seeking behaviour or socio-economic factors |
| Cluster-randomised trials(7,8,21) | Direct and indirect (herd) protection | Captures community-level as well as individual effects; strong internal validity | Expensive; complex logistics; large sample sizes required; usually impractical post-licensure |
| Individually randomised trials(7) | Direct protective efficacy at the individual level | Considered the benchmark for causal inference; high internal validity | Often unethical or unfeasible after licensure; restricted generalisability to routine programme conditions |

Rotavirus vaccines were most frequently evaluated, accounting for 31 studies (48.4%), many conducted through the WHO-coordinated African Rotavirus Surveillance Network across multiple countries in Eastern and Southern Africa(27). Pneumococcal conjugate vaccines were assessed in nine studies (14.1%), most commonly PCV13, with fewer studies evaluating PCV7 and PCV10. Measles-containing vaccines were examined in eight studies (12.5%). Influenza and Haemophilus influenzae type b vaccines were less frequently studied, with five and four studies respectively. Meningococcal vaccines were evaluated in three studies, while polio and cholera vaccines were rarely assessed, each represented by two studies (3.1%).

The geographical distribution of studies revealed notable disparities across sub-Saharan Africa. Southern Africa accounted for nearly half of all studies (n = 30, 45.5%), followed by West Africa with 20 studies (30.3%) and East Africa with 12 studies (18.2%). In contrast, Central Africa (6.1%) and Northwest Africa (1.5%) were significantly underrepresented. In terms of publication period, there was a marked increase in study output over the last decade, with 86% (n = 57) of studies published between 2010 and 2019. Only nine studies (13.6%) were published between 2000 and 2009, reflecting both the acceleration of vaccine introductions in the region and an apparent expansion in regional research capacity.

**Table 1:**
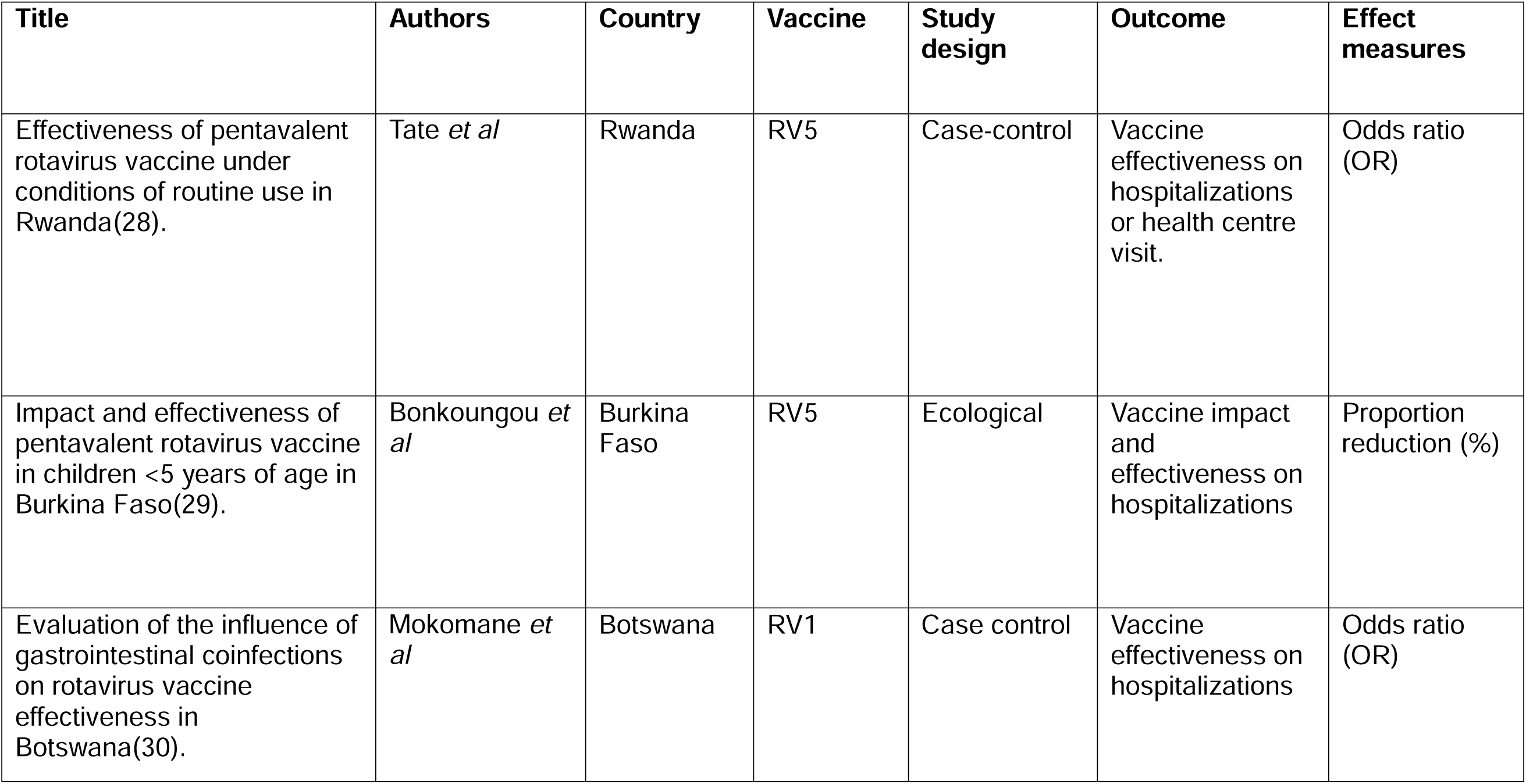
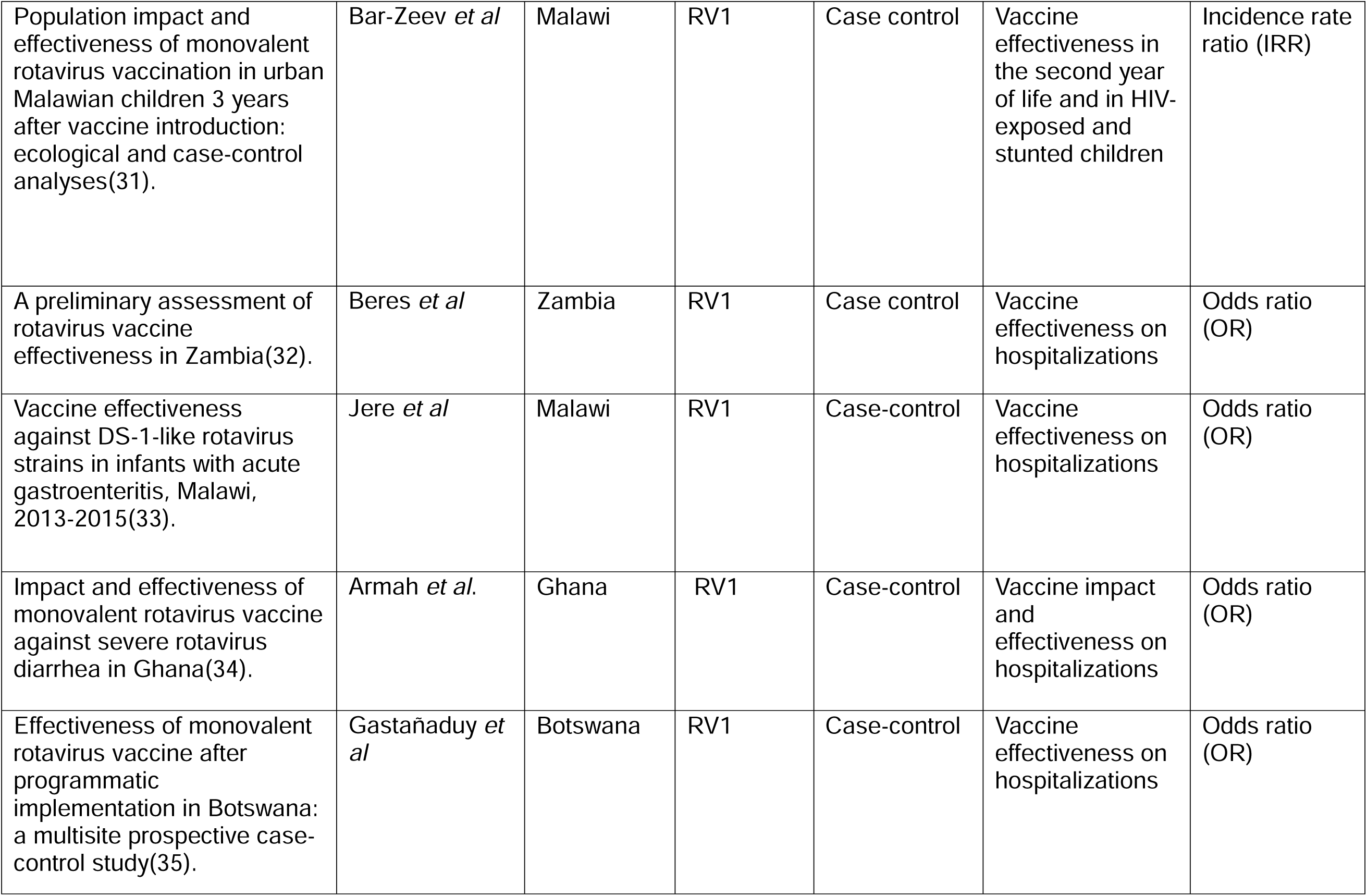
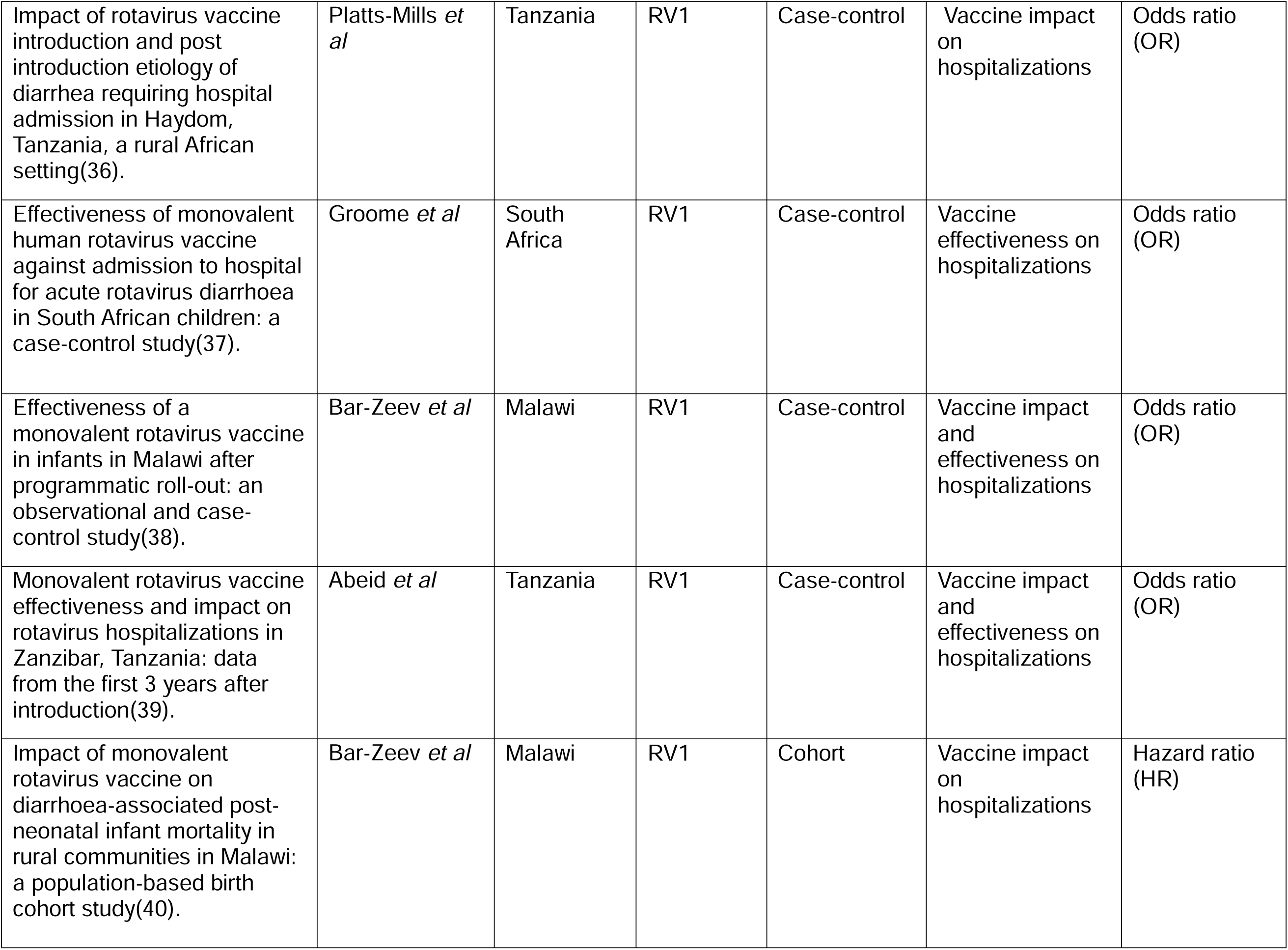
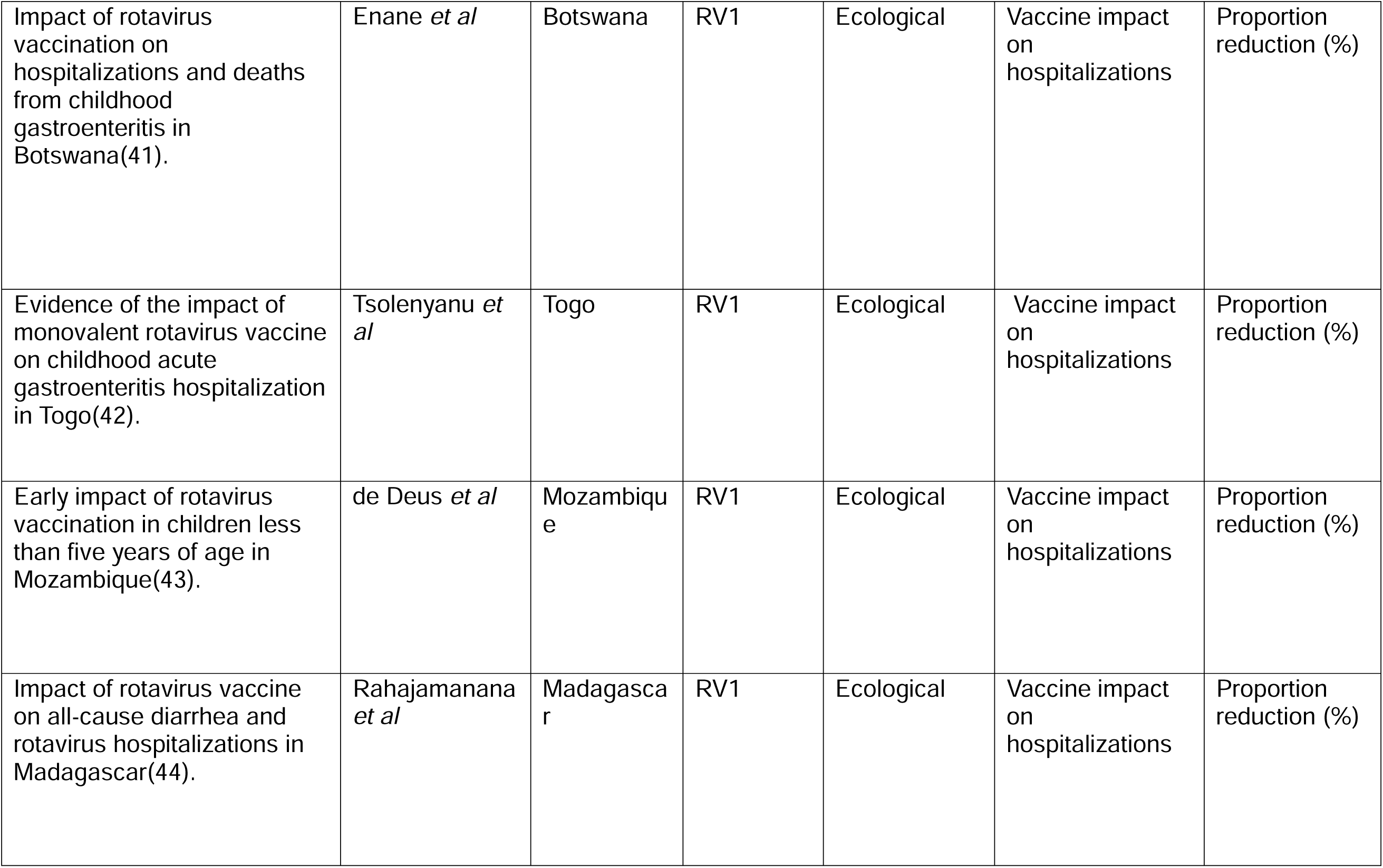
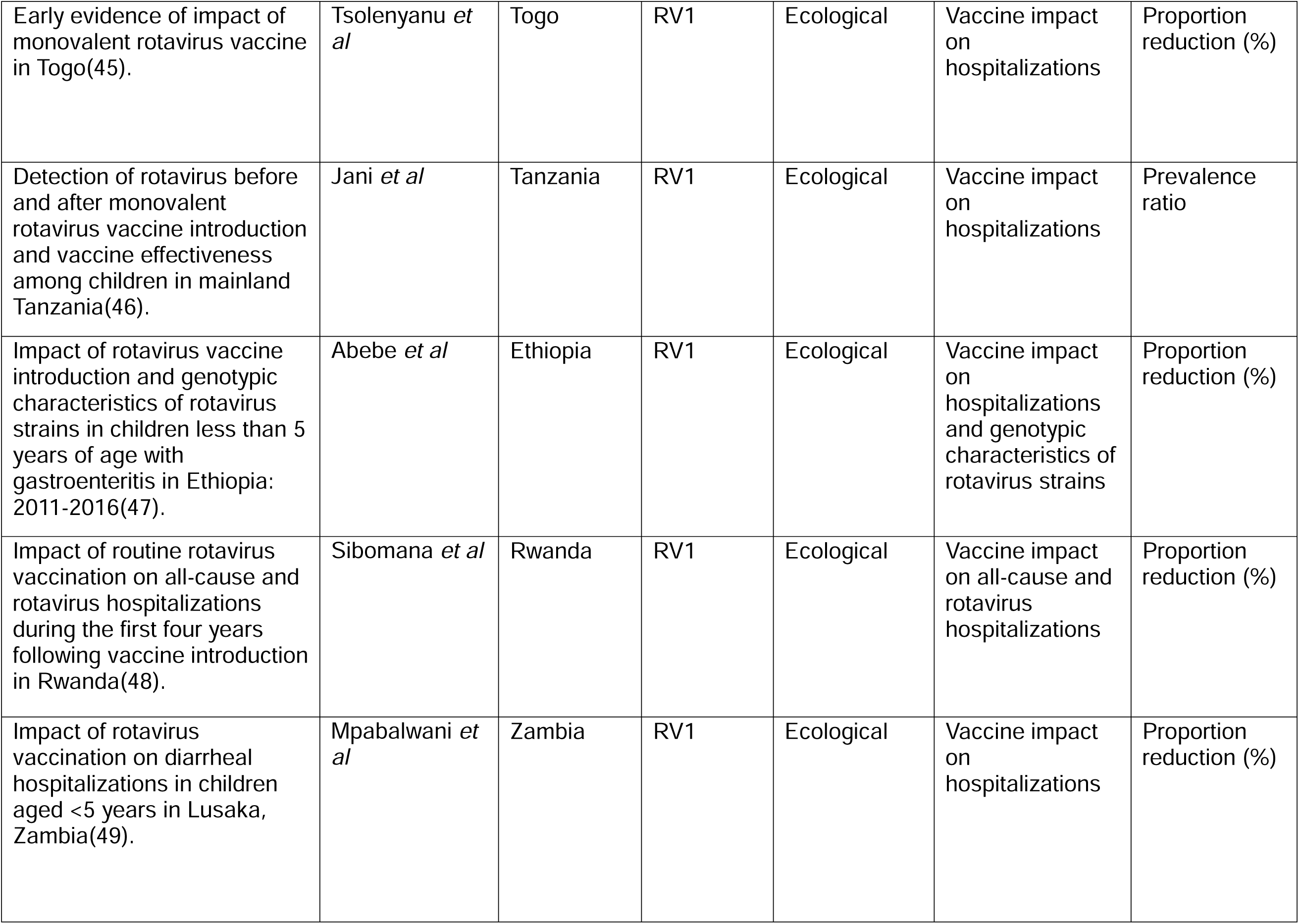
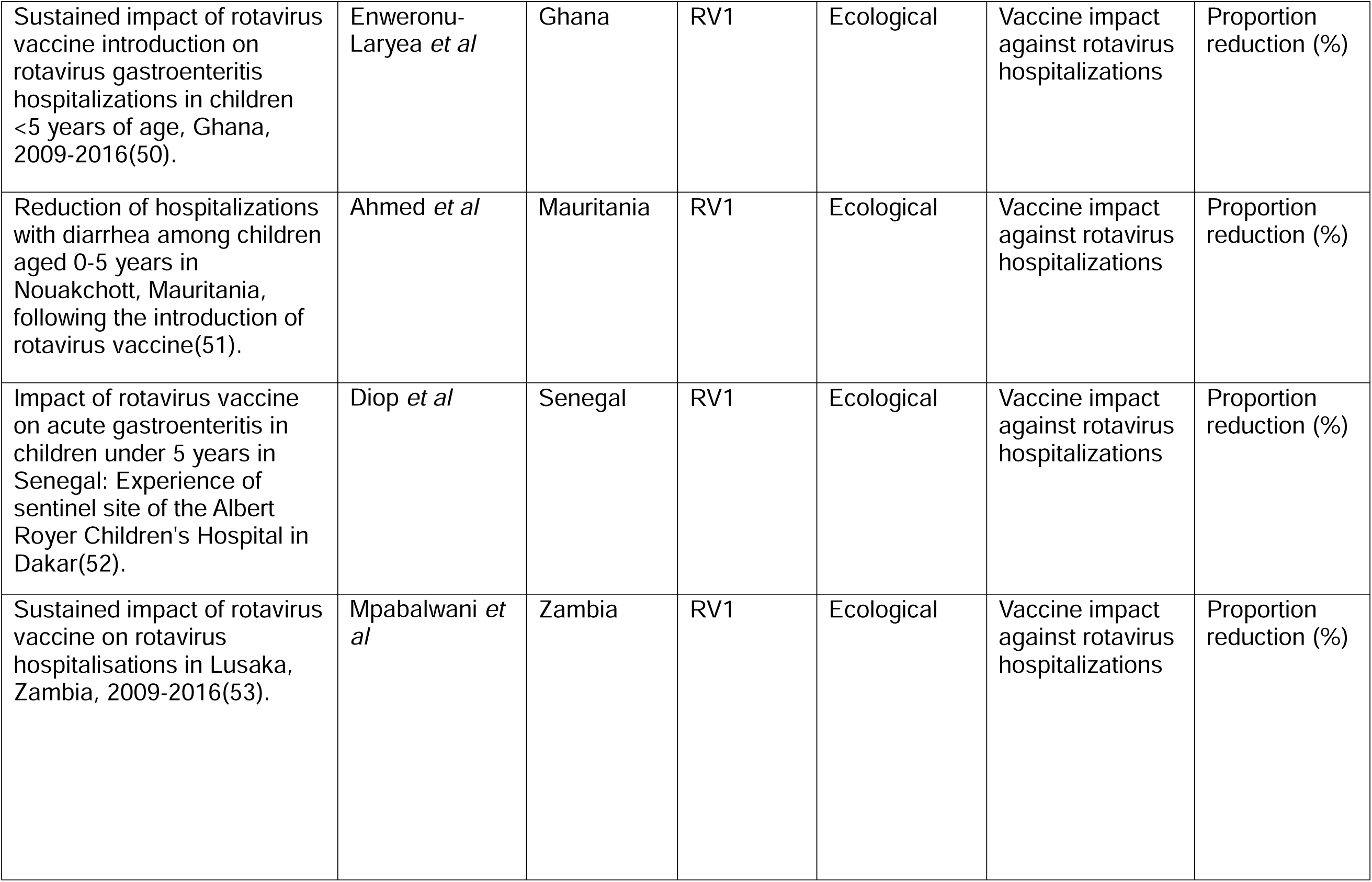
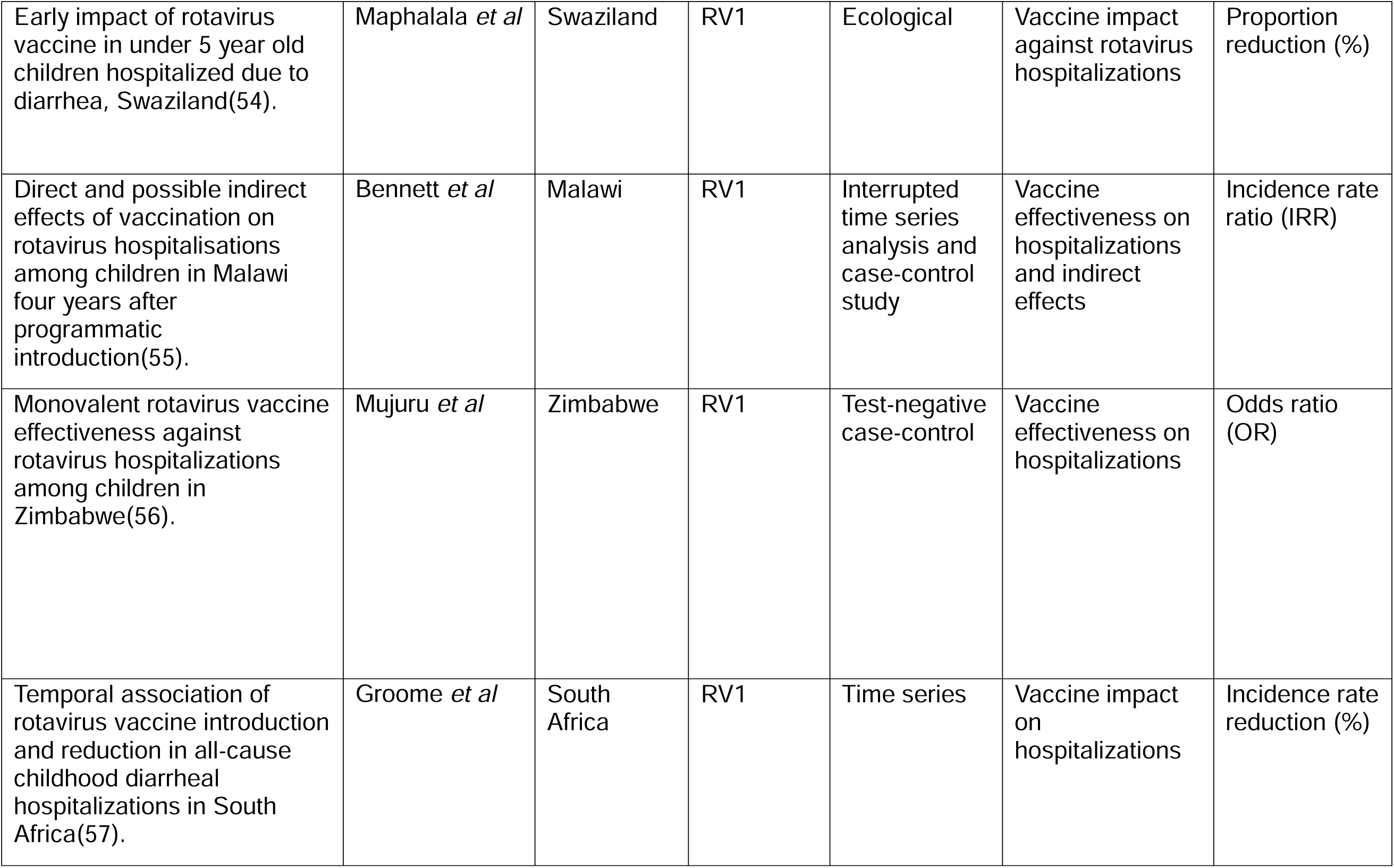
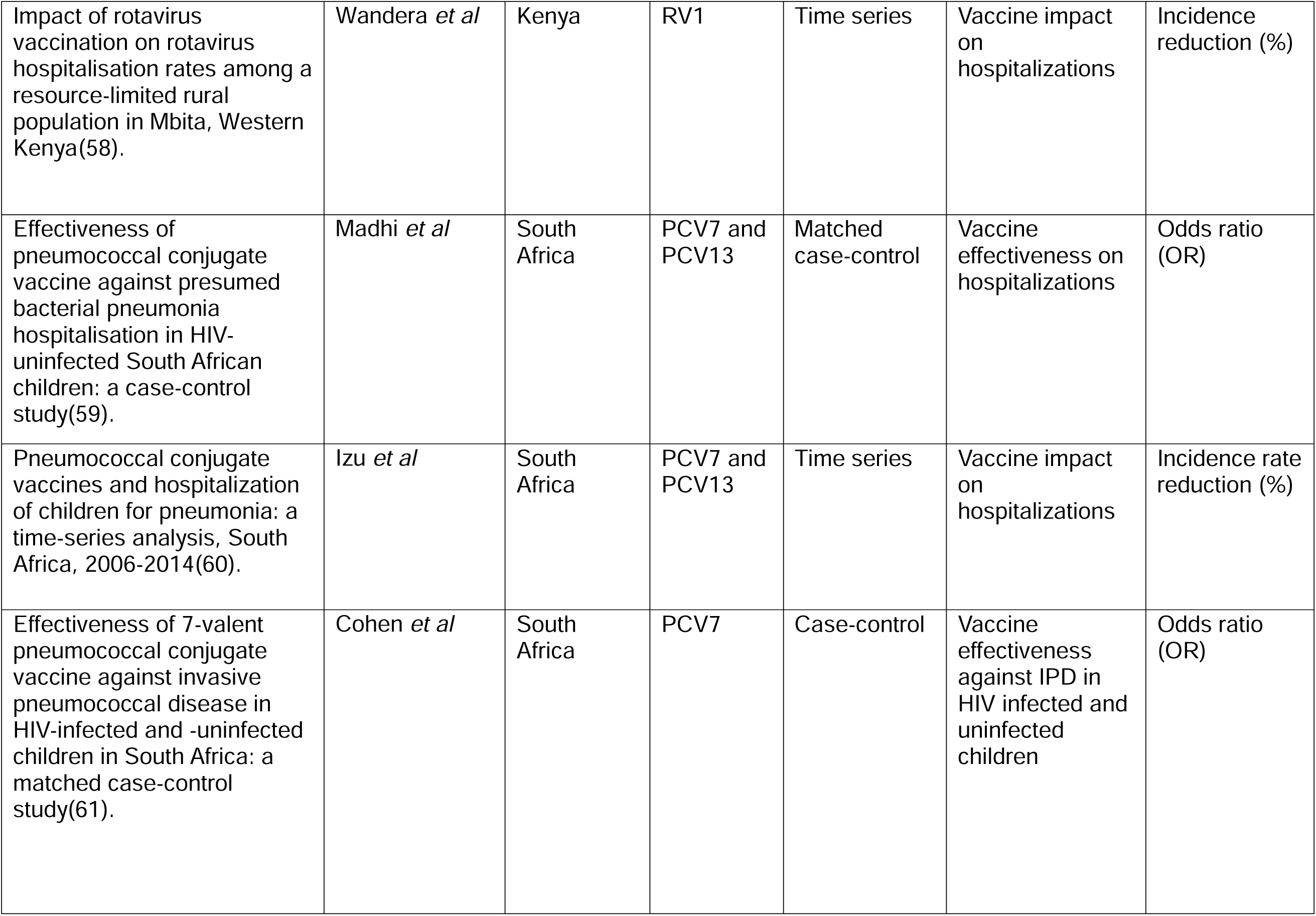
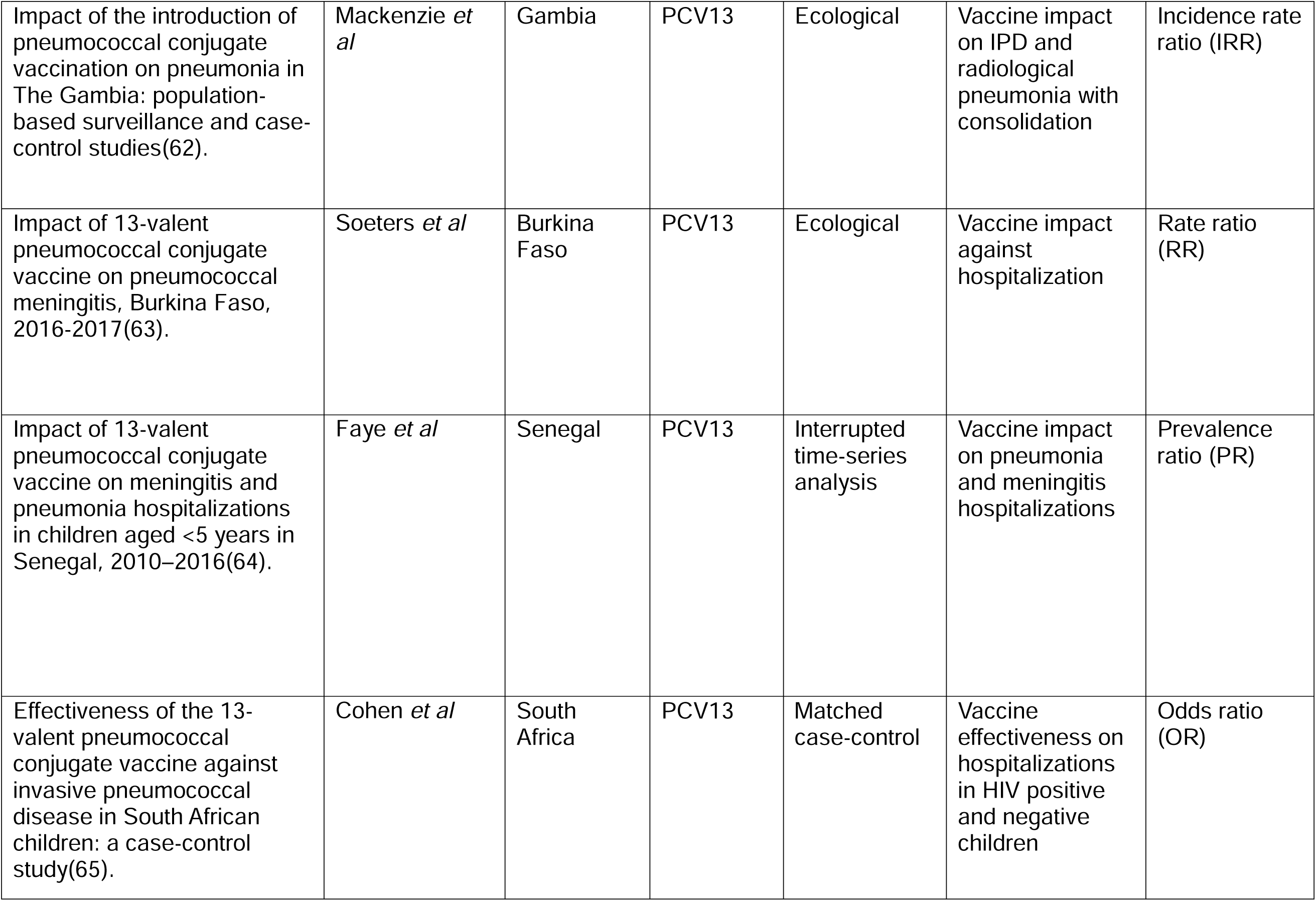
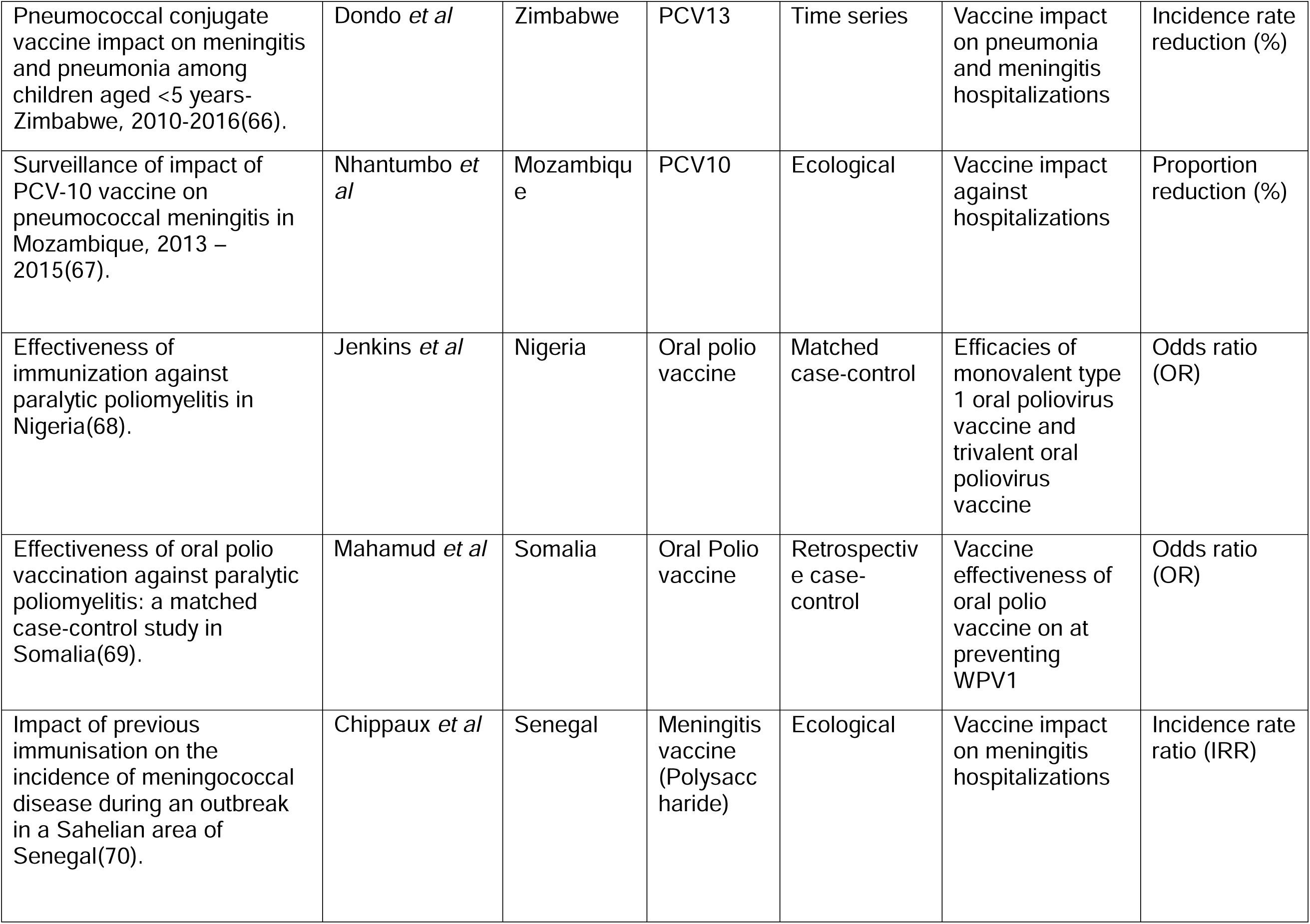
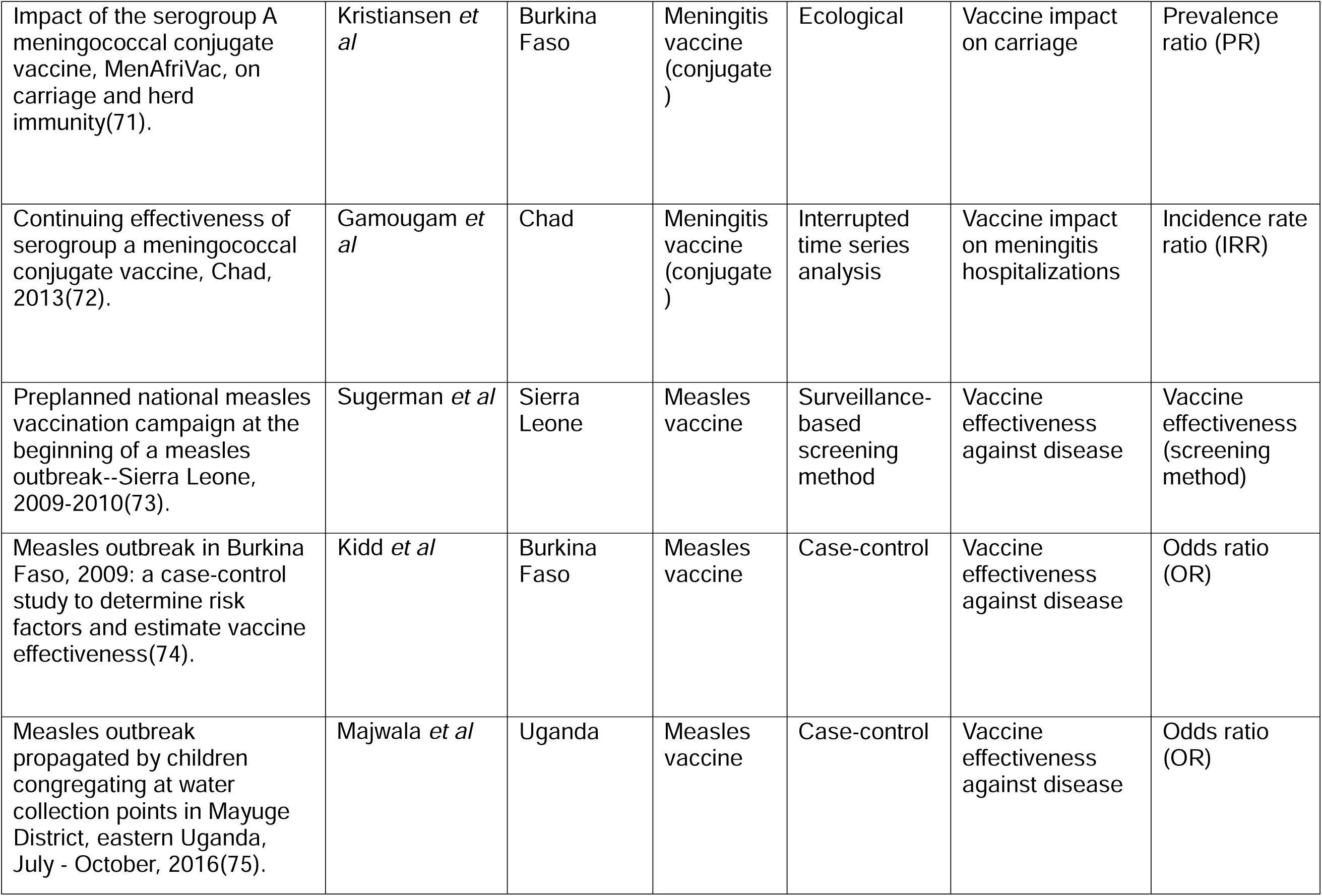
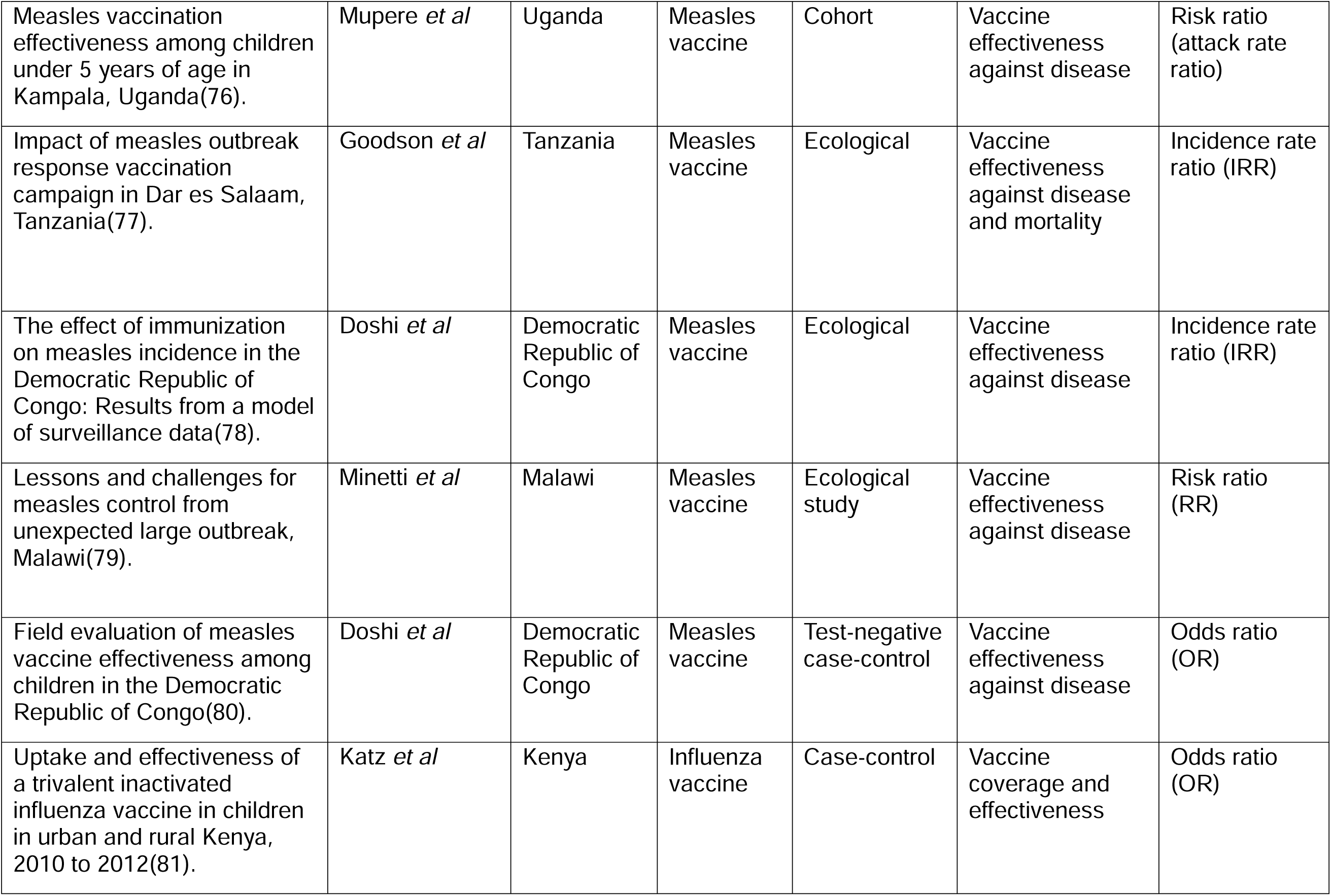
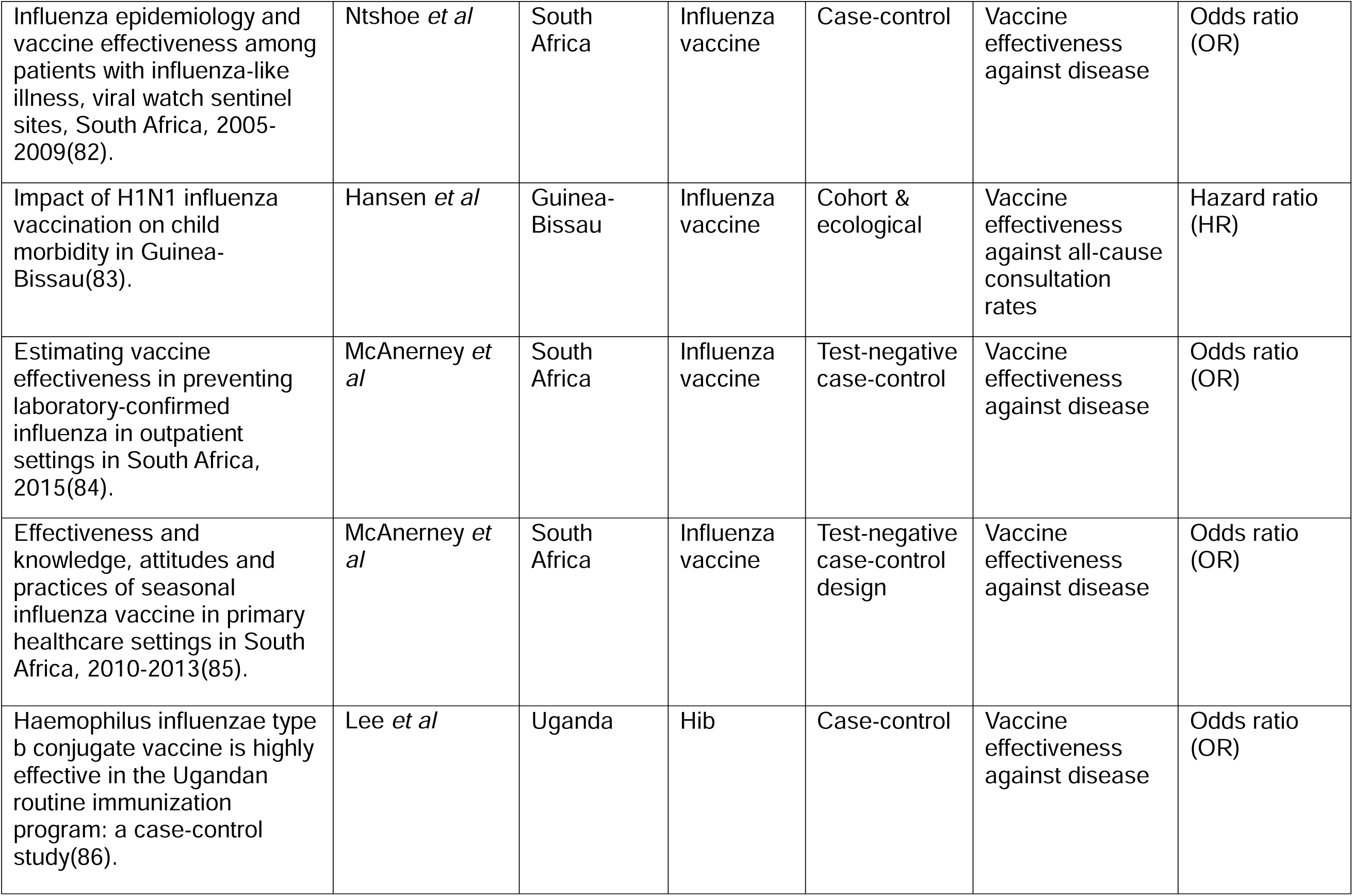
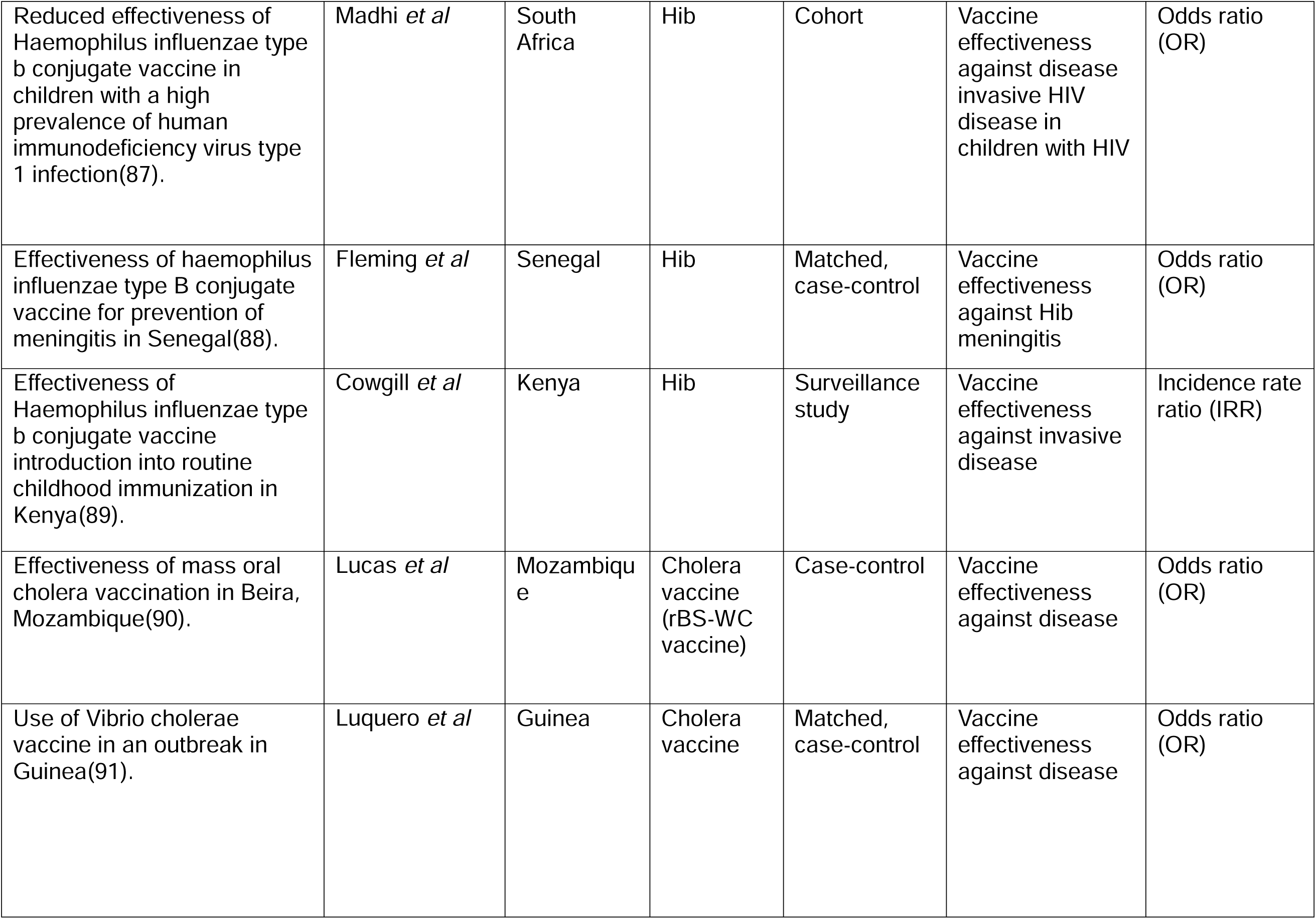
Characteristics of included studies.

Over half of the studies (53%) were funded by non-profit organisations such as WHO AFRO, Gavi, the Vaccine Alliance, and the Bill and Melinda Gates Foundation often in collaboration with public institutions (15%). Public sector funding alone accounted for 12%, while support from the pharmaceutical industry was limited, representing less than 5% of included studies.

**Figure 2:**
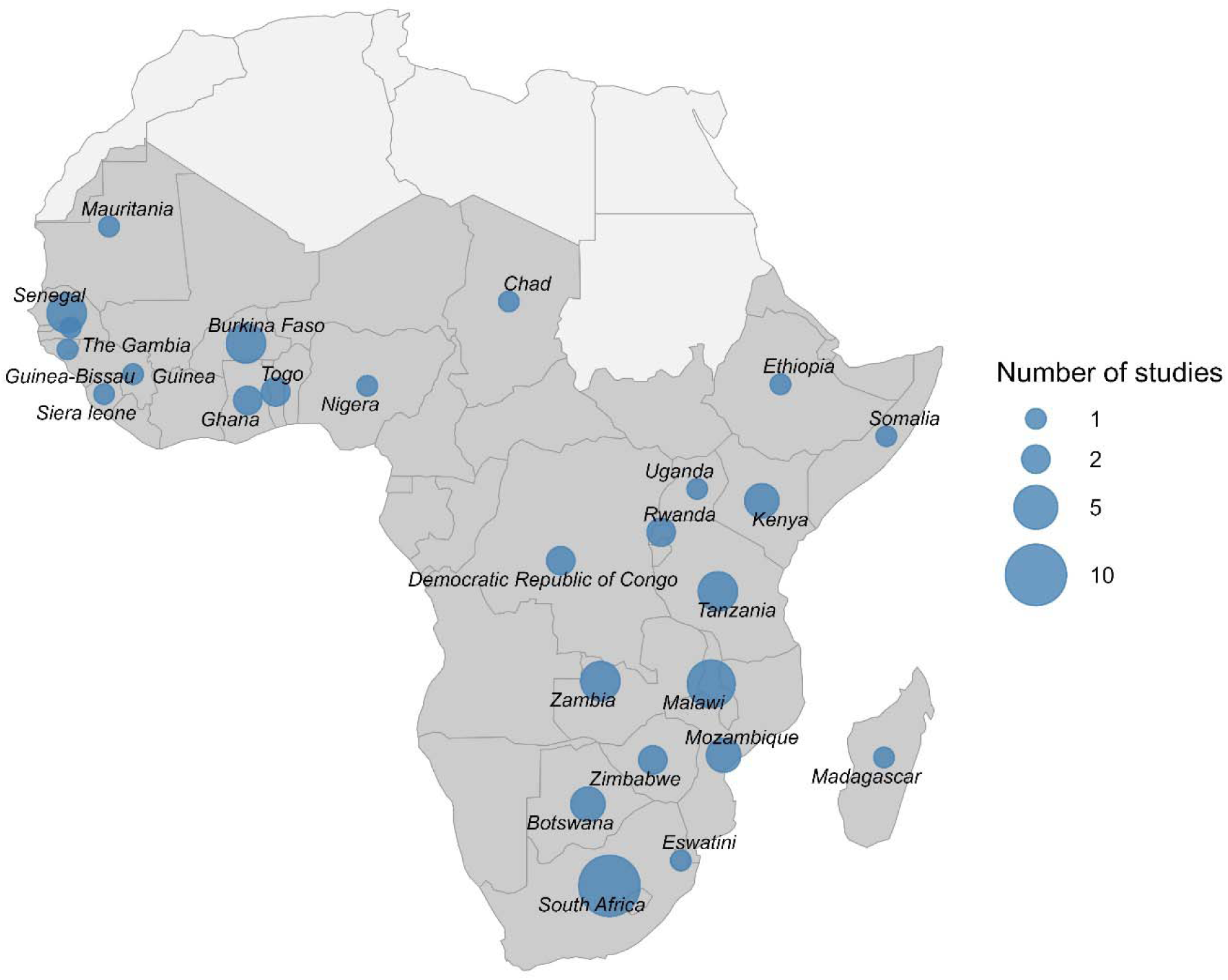
Study locations and number in Sub-Saharan Africa.

**Table 2:**
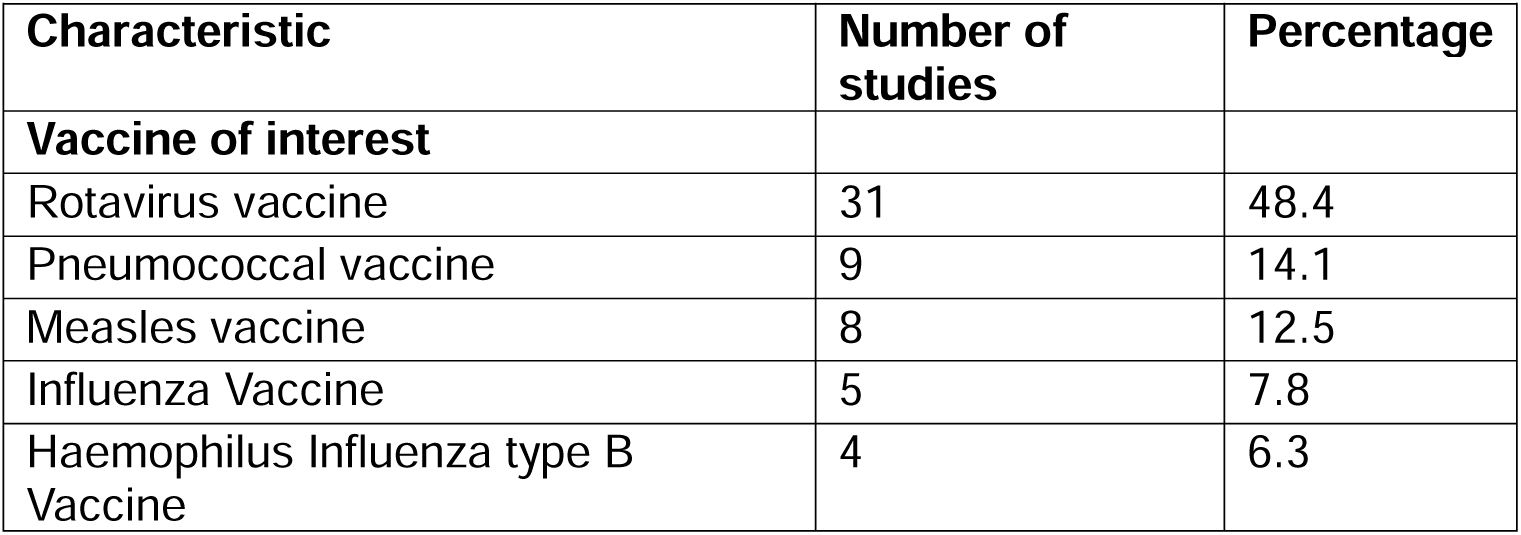
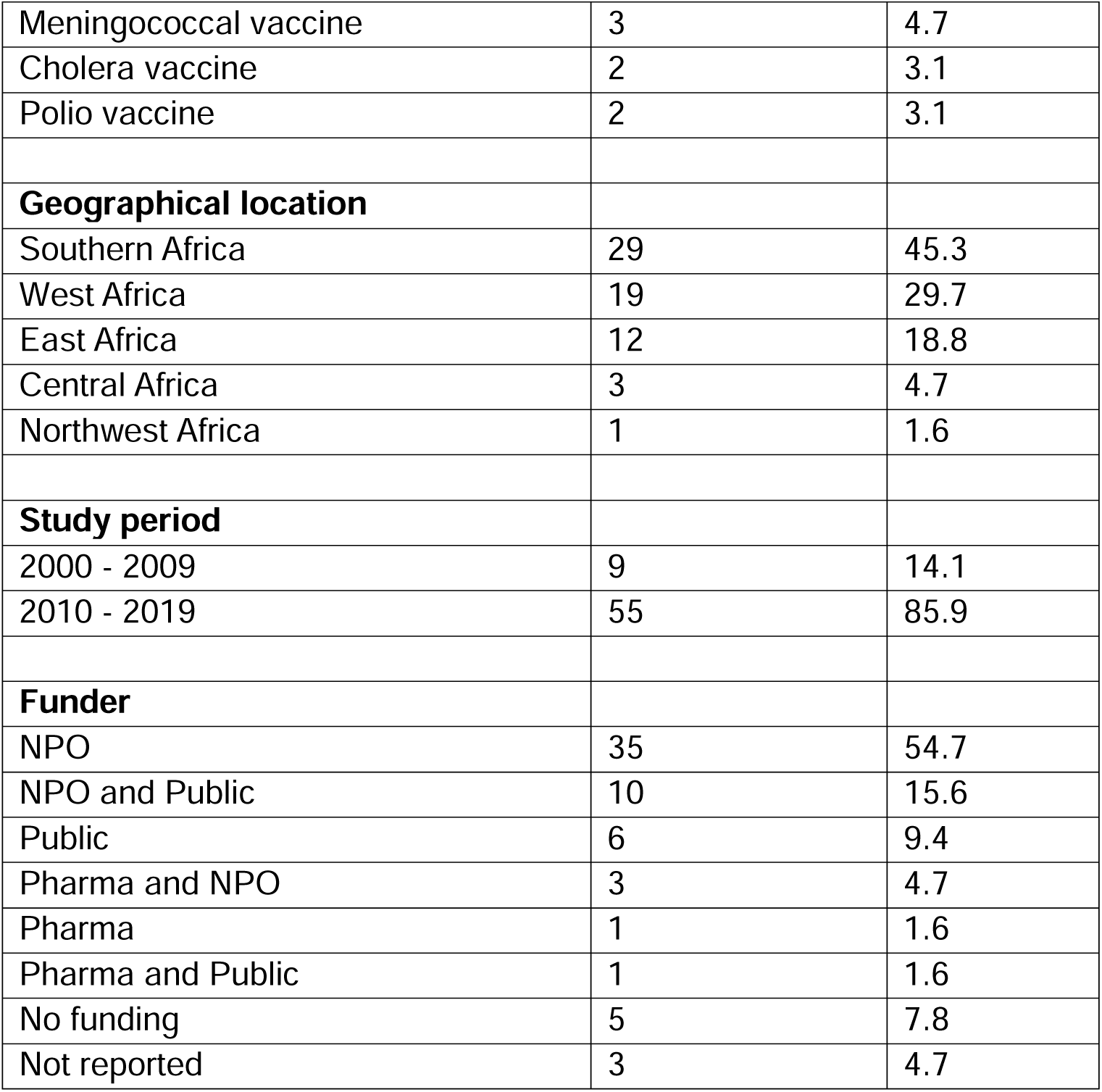
Descriptive statistics of included studies.

| Characteristic | Number of studies | Percentage |
| --- | --- | --- |
| **Vaccine of interest** |  |  |
| Rotavirus vaccine | 31 | 48.4 |
| Pneumococcal vaccine | 9 | 14.1 |
| Measles vaccine | 8 | 12.5 |
| Influenza Vaccine | 5 | 7.8 |
| Haemophilus Influenza type B Vaccine | 4 | 6.3 |
| Meningococcal vaccine | 3 | 4.7 |
| Cholera vaccine | 2 | 3.1 |
| Polio vaccine | 2 | 3.1 |
| **Geographical location** |  |  |
| Southern Africa | 29 | 45.3 |
| West Africa | 19 | 29.7 |
| East Africa | 12 | 18.8 |
| Central Africa | 3 | 4.7 |
| Northwest Africa | 1 | 1.6 |
| **Study period** |  |  |
| 2000 - 2009 | 9 | 14.1 |
| 2010 - 2019 | 55 | 85.9 |
| **Funder** |  |  |
| NPO | 35 | 54.7 |
| NPO and Public | 10 | 15.6 |
| Public | 6 | 9.4 |
| Pharma and NPO | 3 | 4.7 |
| Pharma | 1 | 1.6 |
| Pharma and Public | 1 | 1.6 |
| No funding | 5 | 7.8 |
| Not reported | 3 | 4.7 |

Table 3 summarises the study designs used across the 64 included studies and their consideration of public health and social measures. None of the studies collected or reported data on interventions such as WASH, nutritional improvements, or access to healthcare. Case–control studies were the most common design, comprising 29 studies (45.3%), followed by ecological analyses (n = 24, 37.5%). Neither design accounted for these measures. Time-series approaches were used in seven studies (10.9%), while cohort designs were least represented (n = 4, 6.3%). Two studies combined designs, and classification was based on the primary analytical approach. Although time-series and cohort designs may offer greater scope for causal interpretation, none incorporated information on concurrent public health and social measures. No studies used the screening method or cluster-randomised designs.

**Table 3:**
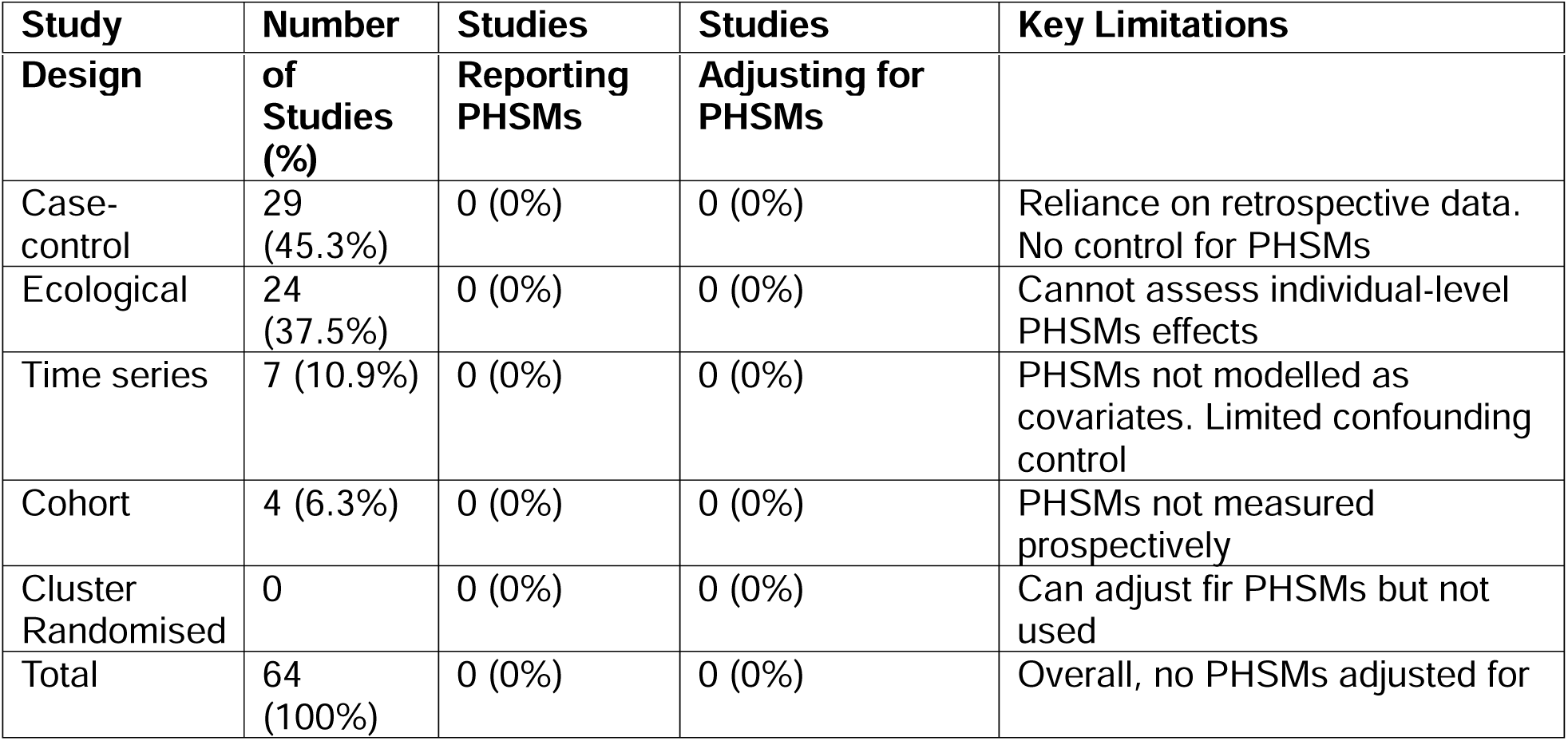
Analysis of study designs and public health and social measures.

| Study | Number | Studies | Studies | Key Limitations |
| --- | --- | --- | --- | --- |

| Design | of Studies (%) | Reporting PHSMs | Adjusting for PHSMs |  |
| --- | --- | --- | --- | --- |
| Case-control | 29 (45.3%) | 0 (0%) | 0 (0%) | Reliance on retrospective data. No control for PHSMs |
| Ecological | 24 (37.5%) | 0 (0%) | 0 (0%) | Cannot assess individual-level PHSMs effects |
| Time series | 7 (10.9%) | 0 (0%) | 0 (0%) | PHSMs not modelled as covariates. Limited confounding control |
| Cohort | 4 (6.3%) | 0 (0%) | 0 (0%) | PHSMs not measured prospectively |
| Cluster Randomised | 0 | 0 (0%) | 0 (0%) | Can adjust for PHSMs but not used |
| Total | 64 (100%) | 0 (0%) | 0 (0%) | Overall, no PHSMs adjusted for |

### Vaccine effectiveness and impact

Rotavirus vaccines reported effect estimates were consistently protective in direction, although the magnitude of both effectiveness and impact varied across study designs, outcome measures and population. Case-control and test-negative studies most often reported VE of 52% – 78% against severe or hospitalised laboratory-confirmed rotavirus(30,32,33,92–96). Ecological studies, including interrupted time-series designs, reported similar declines in diarrhoeal hospitalisations following vaccine introduction with reductions in rotavirus positivity ranging from 35% – 81%(29,45,46,49,52,54,57,97–105). Two studies found no evidence that malnutrition or HIV exposure altered rotavirus vaccine effectiveness(37,106). There was one cohort study that reported a VE of 34% against rotavirus-associated diarrhoea mortality in Malawi(40). These findings suggest that the direction of rotavirus vaccine effects ware consistent across study designs. Differences in reported effectiveness and impact may reflect variation in outcomes and epidemiological context.

Pneumococcal conjugate vaccine studies reported variable effects across outcomes and study designs. Ecological analyses generally indicated substantial reductions in invasive pneumococcal disease and meningitis following vaccine introduction, with reported effectiveness estimates often exceeding 75%(62,67,107). Individual-level case-control studies showed lower effectiveness against broader clinical pneumonia outcomes but higher protection against invasive pneumococcal disease, with estimates ranging from around 57% to 85% depending on HIV status and outcome definition(59,61,65). Time-series analyses also showed heterogeneous population-level impacts. While some settings reported moderate reductions in pneumonia hospitalisations in both HIV infected and uninfected(60), others reported smaller or non-significant changes(108). There were larger declines in meningitis and pneumonia hospitalisations were observed in Zimbabwe (109). Overall, variation in reported PCV effects appears to reflect differences in outcomes, HIV status and vaccine valency.

Other vaccines including measles, influenza, meningococcal, Hib, cholera, and polio vaccines generally showed protective effects. Measles vaccine studies consistently reported high levels of protection across study designs. Case-control evaluations from Uganda estimated vaccine effectiveness of around 74–75%(75,110). A screening method analysis conducted during an outbreak investigation in Sierra Leone reported a similar effectiveness of 74%(111). Ecological analysis from Malawi also indicated strong protection, with effectiveness estimated at 83.9% for a single dose and 90.5% for two-dose schedules. Together these findings indicate that protection from measles vaccination remains strong across different countries in SSA. Estimates were more variable in outbreak settings.

Influenza vaccine studies showed much more variability in effect estimates across seasons and settings. Case-control studies in South Africa reported effectiveness estimates ranging from −14% to 67% across five influenza seasons, with wide confidence intervals in several years(84,112,113). A study from Kenya reported more consistent protection. Vaccine effectiveness ranged from 39% to 57% depending on follow-up duration(81). One cohort analysis from Guinea-Bissau examining non-specific effects found no evidence of reduced consultation rates among vaccinated children and suggested a weaker decline compared with unvaccinated children(83). Together these findings indicate that influenza vaccine effectiveness varied substantially across seasons and contexts, with estimates ranging from negligible or uncertain protection to moderate effectiveness depending on circulating strains, follow-up period and study design.

In contrast, Hib vaccine studies consistently reported high effectiveness against invasive disease and meningitis. Case-control evaluations estimated effectiveness of around 96–99% in Uganda and Senegal(86,88). A cohort study from South Africa reported effectiveness of 83% among fully vaccinated children, with substantially lower protection in HIV-infected children(87). Surveillance data from Kenya also indicated high effectiveness of around 88%(89). Meningococcal vaccine studies reported contrasting findings by vaccine type. Ecological analysis from Senegal evaluating the polysaccharide vaccine found no significant change in meningitis incidence among children under five years(114). In contrast, ecological evaluation of the MenAfriVac conjugate vaccine in Chad reported high effectiveness of approximately 89.6%(115). These findings suggest substantially stronger population-level impact for the conjugate vaccine compared with earlier polysaccharide formulations.

Evidence for cholera vaccines was limited but consistently indicated high protection against disease. Case-control studies evaluating oral cholera vaccines reported effectiveness of 78% for the recombinant cholera toxin B-subunit killed whole-cell vaccine (Dukoral™) in Mozambique and 86.6% for the killed whole-cell vaccine Shanchol™ in Guinea. These findings suggest substantial protection across settings, although the small number of studies limits broader conclusions about variability in vaccine impact across epidemiological contexts(90,91).

Evidence for polio vaccines was limited to two case-control studies but indicated moderate to high effectiveness against paralytic disease. Estimates varied by vaccine formulation and dose number. In Nigeria, monovalent oral polio vaccine (mOPV1) showed substantially higher effectiveness (67%) against type 1 poliovirus than the trivalent formulation (16%). Trivalent vaccine effectiveness remained low (18%)against both type 1 and type 3 disease(68). A matched case-control study from Somalia reported overall effectiveness of around 70%, with higher protection observed among children receiving four or more doses. These findings highlight the importance of vaccine formulation and cumulative dosing in determining observed protection against poliovirus infection(116). As with the other vaccines, none of these studies explicitly measured or adjusted for concurrent public health and social measures, despite many evaluations being conducted in settings undergoing rapid contextual change.

### Heterogeneity in effectiveness estimates: rotavirus case study

For rotavirus, 12 case-control and test-negative studies reported ratio-based effectiveness estimates against hospitalised or severe laboratory-confirmed rotavirus gastroenteritis in children under five. These studies used comparable individual-level designs and outcomes. Effect estimates were consistent across settings. In a random-effects meta-analysis, the pooled odds ratio was 0.42 (95% CI 0.35–0.51). No between-study heterogeneity was detected (I² = 0%, τ² = 0; X² p = 0.67). Several small studies reported wide confidence intervals. This uncertainty reflected limited precision within individual studies rather than differences between studies once design and outcome definitions were aligned.

Variation in post-licensure vaccine effectiveness and impact was not uniform across vaccines. It reflected differences in pathogen biology, outcome specificity, and contextual modifiers(Supplementary Table S1). In settings where public health and social measures change over time yet remain unmeasured, this distinction shapes how vaccine impact evidence should be interpreted and compared across studies.

**Figure 3:**
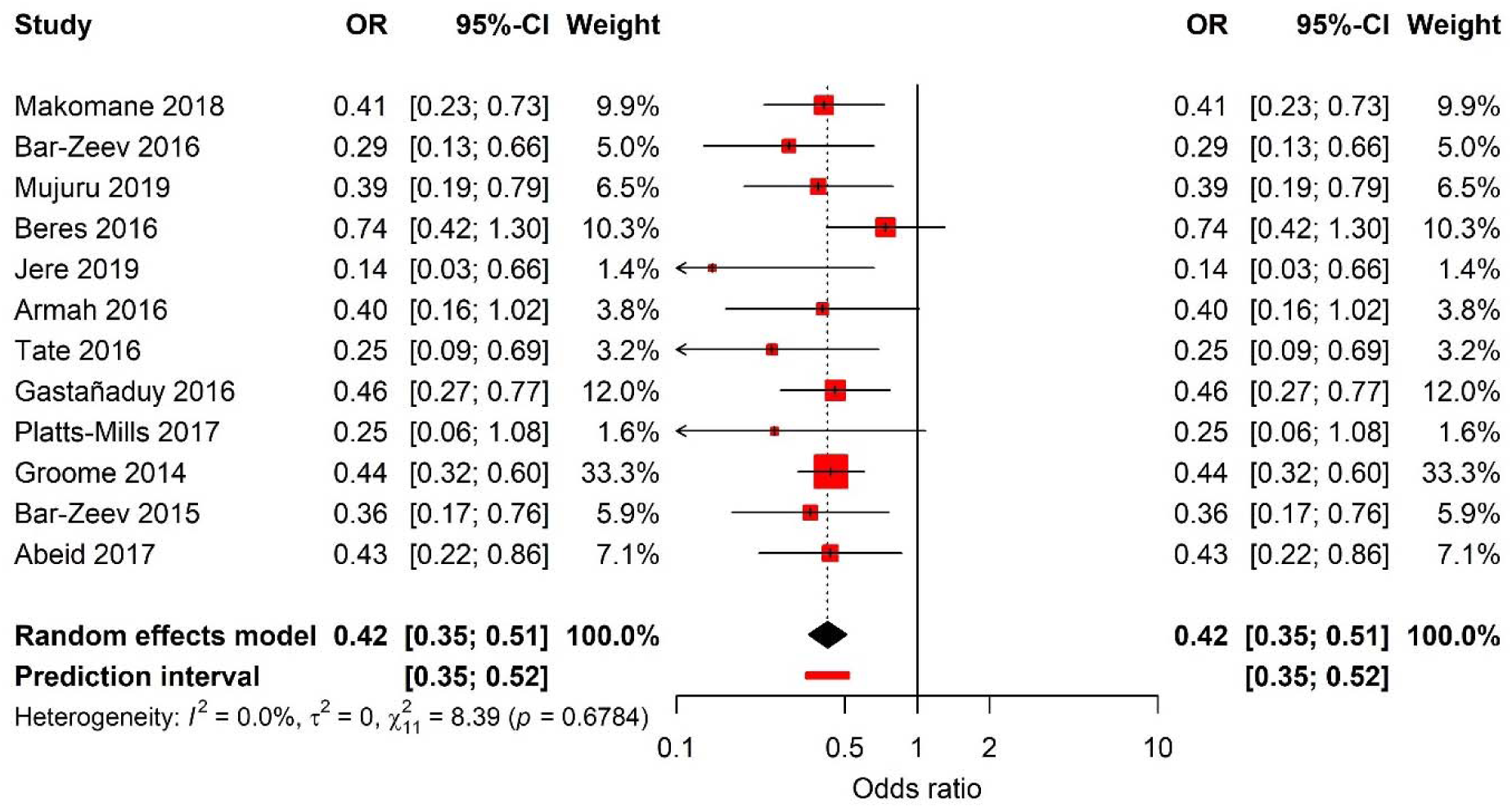
Forest plot of rotavirus vaccine effectiveness from case-control studies in children under five years in sub-Saharan Africa.

## Discussion

### Summary of key findings

To the best of our knowledge, this is the first systematic review to assess the extent to which post-licensure vaccine effectiveness and impact studies conducted in children under five in sub-Saharan Africa account for PHSMs in study design and/or analysis. The key finding of this review is that none of the 64 studies across eight vaccine-preventable diseases reported or adjusted for the effects of PHSMs such as improved WASH practices, nutritional supplementation, or improved healthcare access. This represents a methodological gap in methodology for vaccine evaluations as PHSMs are known to modify vaccine effect estimates (117).

All studies included in this review used observational designs, most commonly case-control and ecological approaches. Case-control studies allow adjustment for individual-level confounders, though this relies on the assumption that cases and controls are comparable with respect to unmeasured exposures or that relevant confounders are adequately captured. This assumption may not hold for PHSMs, which are often not measured at the individual level and may vary over time and across populations. Time-series and cohort designs were less frequently used, which may reflect limited availability of pre-vaccine data, constraints in routine data systems, and the resources required to implement longitudinal studies.

The absence of adjustment for public health and social measures did not have uniform implications across vaccines. In the Results, rotavirus effectiveness estimates from comparable individual-level designs and outcomes were stable across settings, despite the absence of measured PHSMs. Pneumococcal conjugate vaccine estimates varied across outcomes and contexts, even within similar case-control designs. Influenza estimates showed pronounced seasonal and strain-related variation, while Haemophilus influenzae type b and meningococcal conjugate vaccines were associated with large and sustained reductions in disease in high-coverage settings. These patterns suggest that unmeasured PHSMs may influence vaccine effects in ways that differ by pathogen, rather than acting as a uniform source of variability.

### How to adjust for public health and social measures across study designs

Case-control studies collect individual-level data on vaccination status and disease outcomes, which in principle provides a statistical framework that allows adjustment for confounding factors. Their relatively low cost and operational feasibility stems from reliance on retrospective data and their ability to be implemented rapidly(15). They were the most common study design in this review (47%). Interpretation of vaccine effectiveness estimates from case-control studies nevertheless relies on an assumption that cases and controls arise from the same underlying population and experience comparable exposure to contextual interventions that influence disease risk. When exposure to public health and social measures differs between cases and controls, or when these exposures are not measured, residual confounding may arise. As observed in this review, case-control studies did not report or adjust for PHSMs. Data on such measures are often not collected at the individual level and may be prone to recall bias. Their retrospective nature further limits the availability of reliable information on PHSMs that vary over time.

A case-control study in Malawi stratified rotavirus VE by nutritional status and reported substantially higher effectiveness (VE 78.1% [95% CI: 5.6%–94.9%]) in well-nourished children than in stunted children (VE 27.8% [95% CI: –99.5% to 73.9%])(118). This pattern is consistent with nutritional status acting as an effect modifier rather than a confounder. Stratified estimates are therefore informative. The challenge arises when individual-level anthropometric data are not available while nutritional status shifts at the population level over time. Under these conditions, the overall vaccine effect becomes difficult to interpret, as the underlying distribution of nutritional status changes in ways that are not captured in the analysis. This may introduce variability in effectiveness and impact estimates even within individual-level case-control designs. Collection of anthropometric data alongside vaccination status would strengthen future rotavirus evaluations. Ecological analyses would require corresponding adjustment for population-level indicators of nutritional status.

Ecological studies remain one of the most used designs for evaluating vaccines accounting for approximately 36.4% of studies in this review. They compare of disease trends before and after vaccine introduction using population aggregates and are unable to measure individual-level uptake or exposure to PHSMs(11). They report disease trends from routine surveillance data which do not include data on PHSMs, and if they do, it is incomplete and inconsistently reported. Vaccine introductions in sub-Saharan Africa are usually accompanied by introduction of PHSMs such was WASH, bed nets etc(119). Without granular data, it is impossible to disentangle the effects of PHSMs which potentially leads to overestimation or underestimation of effect estimates. Additionally, the temporal sequence of changes in PHSMs relative to vaccine introduction is often unclear, further complicating causal inference.

Cohort studies offer very strong methodological advantage for adjusting for PHSMs. They collect exposure data (vaccination status) at baseline and follow-up individuals over time allowing comparison of disease incidence between vaccinated and unvaccinated children(120). Compared to ecological designs they provide individual-level data ideal for adjusting for time-varying PHSMS confounders. For instance, data on improved PHSMs access in a particular township mid-study can be collected and adjusted for accordingly. Despite this potential, none of the four cohort studies in this review adjusted for PHSMs which is likely because of the lack of lack data on PHSMs source, nutrition and hygiene behaviours. This can lead to residual confounding, particularly when PHSMs such as WASH are unequally distributed between vaccinated and unvaccinated children. Moreover, cohort studies may suffer from confounding by indication, as children who receive vaccines may differ systematically from those who do not in terms of maternal education, health-seeking behaviours, and other factors linked to PHSMS exposure.

Interrupted time series (ITS) analysis are quasi-experimental designs that assess vaccine impact using aggregated surveillance data across multiple time points. These models can adjust for trend and seasonality to provide vaccine effect estimates following vaccine introduction. However, despite this potential, ITS rarely adjust for time-varying PHSMs due to data limitations or methodological constraints which introduces confounding. For instance, A South African pneumococcal time-series study showed 33% (50% credible interval, CrI: 6 to 52) reduction in pneumonia hospitalisations post-introduction of pneumococcal conjugate vaccines but lacked data on coincident improvements in HIV treatment access from 2006 to 2014(121). Controlled ITS have a control group which allows researchers to control for time-varying PHSMs for vaccine impact estimates(13). There is a need for researchers to link time series data to demographic health systems data to enhance the ability of ITS to control for co-occurring interventions and strengthen impact estimates.

### Recommendations for future studies

Future post-licensure vaccine effectiveness and impact studies in sub-Saharan Africa should prioritise improved measurement of PHSMs that plausibly modify vaccine effects and change over short time scales. Case-control studies should collect individual-level data on nutrition, WASH exposure, malaria prevention, and other contextually relevant factors that are known to influence susceptibility or immune response. These variables should be incorporated for matching in design or in analytical models to allow estimation of effect modification rather than treated solely as sources of confounding.

Matching or adjustment for broader factors such on geography, seasonality may provide partial control for contextual variation and can be considered as complementary strategies. However, these approaches do not replace direct measurement of rapidly evolving PHSMs. In ecological and time-series evaluations, strengthening routine data systems such as District Health Information Systems is crucial for capturing population-level indicators of nutrition, WASH and other key interventions, enabling better interpretation of vaccine impact alongside changes risk factors. As climate variability affects disease seasonality and risk, improving the capture of environmental and social indicators will become more important.

Cohort studies should prioritise the collection of time-varying PHSMs data and adjust for PHSMs accordingly. For interrupted time-series analyses, linkage aggregated health data with routine sources such as WASH databases or HIV programme records can improve adjustment for concurrent interventions and reduce residual confounding. Ministries of Health should be supported to strengthen surveillance platforms incorporating indicators of PHSMs into routine health information systems to improve monitoring and vaccine impact assessment. Where PHSMs change rapidly and are not captured at the individual level, alternative evaluation designs may be required.

One such approach is cluster randomised evaluation designs as used in the RTS,S/AS01 malaria vaccine pilot implementation and evaluation (MVIE) in Malawi, Ghana and Kenya (122). This allowed unmeasured contextual factors such as health system strengthening, care-seeking behaviour, and concurrent malaria interventions such as bed net usage and indoor residual spraying to be balanced across study arms by design. While this design doesn’t eliminate effect modification at the individual level, it allows estimation of population-average treatment effects that are less sensitive to confounding by time-varying and unmeasured PHSMs. While cluster randomised post-licensure evaluations are not routinely feasible, the RTS,S experience illustrates that such designs can be justified where safety, effectiveness, and programme impact need to be assessed under real-world conditions and where conventional observational designs are likely to yield unstable estimates.

### Implications for policy and practice

Design limitations can lead to underestimation or overestimation of vaccine effect estimates. This has implications for immunisation programmes in low-resource settings, where competing priorities for limited resources may lead to missed opportunities to identify and prioritise synergistic interventions that could enhance the overall impact of vaccination. For example, governments could combine immunisation campaigns with nutrition interventions or distribution of insecticide treated bed nets. For diarrhoeal disease such as cholera, reactive vaccination campaigns should be implemented alongside WASH interventions. It is crucial to collect and adjust for data on PHSMs implemented alongside vaccination programs during evaluations.

Neglecting PHSMs may lead health authorities in LMIC settings to place excessive emphasis on vaccination alone, sidelining complementary interventions such as nutrition programmes, WASH improvements, and maternal health education. At the same time, underestimation of vaccine effects may result in deprioritising effective vaccines, risking robust coverage and outbreak control. Vaccination remains among the most cost-effective public health interventions across all settings(123). Delaying or abandoning vaccine introduction based on robust effect estimates may increase costs for both households and health systems while leaving populations vulnerable to preventable disease.

Underestimated vaccine impact may undermine public and political confidence in vaccines, especially in the face of rising vaccine hesitancy as experienced with COVID-19 vaccines(124). Adjusting for PHSMs for post-licensure studies is also an opportunity for LMICs to expand national data platforms like DHIS to routinely capture PHSMs indicators. Health facility registers and survey questionnaires could be updated to include nutrition, WASH, and other co-interventions to improve evidence quality and support integrated programme planning.

### Strengths and limitations

The review employed a comprehensive and reproducible search strategy across multiple databases including PubMed Central, EMBASE, MEDLINE, and CINAHL and adhered to PRISMA guidelines. We included a wide range of observational study designs, funding sources and vaccines which allowed for a comprehensive appraisal of the methodological approaches employed in post-licensure vaccine evaluations. Furthermore, our dual-reviewer approach to screening, extraction, and quality assessment enhanced the rigour and reliability of the review process.

However, our systematic review had several limitations. The review was restricted to English-language publications, which may have led to the exclusion of relevant studies conducted in francophone and lusophone countries within the region which also have strong immunisation programmes. The review was also limited to countries in sub-Saharan Africa, which excludes evidence from other regions and may limit the generalisability of the findings beyond this context. We relied on published data where analyses were constrained by what was reported in the articles. It is possible that some researchers measured PHSMs but simply did not report them. Finally, we limited our review to 2019 which excluded the evaluation of COVID-19 vaccines.

This scope was chosen to focus on post-licensure evaluation under routine immunisation conditions. COVID-19 vaccine studies were conducted in a markedly different context, with rapidly changing and large-scale PHSMs introduced alongside vaccine rollout. These features create strong co-interventions and secular shifts that are not comparable with the settings in which most established childhood vaccines are evaluated. The rapid expansion of COVID-19–related publications would also have dominated the evidence base and shifted our review away from its primary objectives. Despite these limitations, our review highlights a critical methodological gap in post-licensure vaccine studies and underscores the urgent need for future evaluations to systematically account for PHSMs in both design and analysis.

## Conclusion

This systematic review highlights a critical and under-addressed gap in post-licensure vaccine effectiveness and impact studies conducted in children under five years in sub-Saharan Africa which is the lack of adjustment for PHSMs such as improved nutrition, WASH, malaria control, and HIV-related services. None of the 64 studies included in this review collected, reported, or adjusted for PHSMs in their analysis despite the known confounding effects of these interventions on disease burden and health outcomes. With increasing number of vaccines and broader demands on health budgets it is necessary to provide the most accurate assessments of vaccine contributions to public health in rapidly changing settings.

It is critical to improve the design and reporting of post-licensure vaccine studies in LMIC settings. This includes routine collection of PHSMs-related data, employment of stepped-wedge cluster randomised design and incorporation of such variables into multivariate regression models. Additionally, national health information systems should be strengthened to facilitate the routine capture of relevant PHSMs by researchers. Integrating these improvements into surveillance and evaluation frameworks will improve accuracy and provide context-specific evidence to guide immunisation policies and maximise public health gains from vaccination programmes.

## Data Availability

All data produced in the present study are available upon reasonable request to the authors

## References

1. Table 2 Summary of WHO Position Papers – Recommended Routine Immunizations for Children [Internet]. [cited 2025 Jul 26]. Available from: https://www.who.int/publications/m/item/table-2-summary-of-who-position-papers-recommended-routine-immunizations-for-children

2. Mwenda JM, Parashar UD, Cohen AL, Tate JE. Impact of rotavirus vaccines in Sub-Saharan African countries. Vaccine [Internet]. 2018;36(47):7119–23. Available from: 10.1016/j.vaccine.2018.06.026

3. Ngocho JS, Magoma B, Olomi GA, Mahande MJ, Msuya SE, de Jonge MI, et al. Effectiveness of pneumococcal conjugate vaccines against invasive pneumococcal disease among children under five years of age in Africa: A systematic review. PLoS One [Internet]. 2019 Feb 1 [cited 2026 Mar 13];14(2):e0212295. Available from: https://journals.plos.org/plosone/article?id=10.1371/journal.pone.0212295

4. WHO. Immunization Agenda 2030. 2021;1–24. Available from: https://www.who.int/immunization/ia2030_Draft_One_English.pdf?ua=1

5. Higgins JPT, Soares-Weiser K, López-López JA, Kakourou A, Chaplin K, Christensen H, et al. Association of BCG, DTP, and measles containing vaccines with childhood mortality: systematic review. BMJ [Internet]. 2016 Oct 13 [cited 2025 Jul 28];355:5170. Available from: https://www.bmj.com/content/355/bmj.i5170

6. Singh K, Mehta S. The clinical development process for a novel preventive vaccine: An overview. J Postgrad Med [Internet]. 2016 Jan 1 [cited 2022 Nov 22];62(1):4. Available from: /pmc/articles/PMC4944327/

7. Kahn R, Villar SS, Dean NE, Lipsitch M. Vaccine Trial Designs. Princ Pract Emerg Res Response [Internet]. 2024 Aug 31 [cited 2026 Mar 13];585–610. Available from: https://www.ncbi.nlm.nih.gov/books/NBK614024/

8. Hemming K, Taljaard M. Key considerations for designing, conducting and analysing a cluster randomized trial. Int J Epidemiol [Internet]. 2023 Oct 5 [cited 2026 Mar 13];52(5):1648–58. Available from: 10.1093/ije/dyad064

9. Hanquet G, Valenciano M, Simondon F, Moren A. Vaccine effects and impact of vaccination programmes in post-licensure studies. Vaccine [Internet]. 2013 Nov 19 [cited 2026 Mar 13];31(48):5634–42. Available from: 10.1093/oxfordjournals.epirev.a017931

10. Elizabeth Halloran M, Struchiner CJ, Longini IM. Study Designs for Evaluating Different Efficacy and Effectiveness Aspects of Vaccines. Am J Epidemiol [Internet]. 1997 Nov 15 [cited 2026 Feb 27];146(10):789–803. Available from: 10.1093/oxfordjournals.aje.a009196

11. Morgenstern H. Ecologic studies in epidemiology: Concepts, principles, and methods. Annu Rev Public Health. 1995;16:61–81.

12. Bernal JL, Cummins S, Gasparrini A. The use of controls in interrupted time series studies of public health interventions. Int J Epidemiol [Internet]. 2018 Dec 1 [cited 2023 Oct 24];47(6):2082–93. Available from: 10.1093/ije/dyy135

13. Bernal JL, Cummins S, Gasparrini A. Interrupted time series regression for the evaluation of public health interventions: a tutorial. Int J Epidemiol [Internet]. 2017 Feb 1 [cited 2024 Oct 31];46(1):348–55. Available from: https://pubmed.ncbi.nlm.nih.gov/27283160/

14. Lopez Bernal J, Soumerai S, Gasparrini A. A methodological framework for model selection in interrupted time series studies. J Clin Epidemiol [Internet]. 2018;103:82–91. Available from: 10.1016/j.jclinepi.2018.05.026

15. Orenstein WA, Bernier RH, Dondero TJ, Hinman AR, Marks JS, Bart KJ, et al. Field evaluation of vaccine efficacy. Bull World Health Organ [Internet]. 1985 [cited 2026 Jan 14];63(6):1055. Available from: https://pmc.ncbi.nlm.nih.gov/articles/PMC2536484/

16. Pearce N. Analysis of matched case-control studies. BMJ [Internet]. 2016 Feb 25 [cited 2026 Mar 13];352. Available from: https://www.bmj.com/content/352/bmj.i969

17. Pearce N, Checkoway H. Cohort Studies. Encycl Quant Risk Anal Assess Melnick/Risk [Internet]. 2008 Jan 1 [cited 2026 Mar 13];1–12. Available from: 10.1002/9780470061596.risk0598

18. Wang X, Kattan MW. Cohort Studies: Design, Analysis, and Reporting. Chest [Internet]. 2020 Jul 1 [cited 2026 Mar 13];158(1):S72–8. Available from: 10.1089/jpm.2008.9690

19. Capili B, Anastasi JK. Overview: Cohort Study Designs. Am J Nurs [Internet]. 2021 Dec 1 [cited 2026 Mar 13];121(12):45. Available from: https://pmc.ncbi.nlm.nih.gov/articles/PMC9536647/

20. Praet N, Asante KP, Bozonnat M-C, Akité EJ, Ansah PO, Baril L, et al. Assessing the safety, impact and effectiveness of RTS,S/AS01(E) malaria vaccine following its introduction in three sub-Saharan African countries: methodological approaches and study set-up. Malar J. 2022 Apr;21(1):132.

21. Khanam F, Kim DR, Liu X, Voysey M, Pitzer VE, Zaman K, et al. Assessment of vaccine herd protection in a cluster-randomised trial of Vi conjugate vaccine against typhoid fever: results of further analysis. eClinicalMedicine [Internet]. 2023 Apr 1 [cited 2026 Mar 13];58:101925. Available from: https://www.thelancet.com/action/showFullText?pii=S2589537023001025

22. Patel MM, Steele D, Gentsch JR, Wecker J, Glass RI, Parashar UD. Real-world impact of rotavirus vaccination. Pediatr Infect Dis J. 2011;30(SUPPL. 1):1–5.

23. Bar-zeev N, Tate JE, Pecenka C, Chikafa J, Mvula H, Wachepa R, et al. Cost-Effectiveness of Monovalent Rotavirus Vaccination of Infants in MalawilJ: A Postintroduction Analysis Using Individual Patient – Level Costing Data. 2016;62(Suppl 2).

24. Patriarca PA, Wright PF, John TJ. Factors Affecting the Immunogenicity of Oral Poliovirus Vaccine in Developing Countries: Review. Rev Infect Dis [Internet]. 1991 Sep 1 [cited 2025 Aug 19];13(5):926–39. Available from: 10.1093/clinids/13.5.926

25. Fine PEM. Variation in protection by BCG: implications of and for heterologous immunity. Lancet [Internet]. 1995 Nov 18 [cited 2025 Aug 19];346(8986):1339–45. Available from: https://www.thelancet.com/action/showFullText?pii=S0140673695923489

26. Rayyan: AI-Powered Systematic Review Management Platform [Internet]. [cited 2026 Mar 30]. Available from: https://www.rayyan.ai/

27. Mwenda JM, Tate JE, Parashar UD, Mihigo R, Agócs M, Serhan F, et al. African Rotavirus Surveillance Network – A Brief Overview. Pediatr Infect Dis J [Internet]. 2014 [cited 2026 Mar 30];33(Suppl 1):S6. Available from: https://pmc.ncbi.nlm.nih.gov/articles/PMC12046586/

28. Tate JE, Ngabo F, Donnen P, Gatera M, Uwimana J, Rugambwa C, et al. Effectiveness of Pentavalent Rotavirus Vaccine Under Conditions of Routine Use in Rwanda. Clin Infect Dis an Off Publ Infect Dis Soc Am. 2016 May;62 Suppl 2:S208–12.

29. Bonkoungou IJO, Aliabadi N, Leshem E, Kam M, Nezien D, Drabo MK, et al. Impact and effectiveness of pentavalent rotavirus vaccine in children. Vaccine [Internet]. 2018 Nov;36(47):7170–8. Available from: https://pubmed.ncbi.nlm.nih.gov/29290478/

30. Mokomane M, Tate JE, Steenhoff AP, Esona MD, Bowen MD, Lechiile K, et al. Evaluation of the Influence of Gastrointestinal Co-Infections on Rotavirus Vaccine Effectiveness in Botswana. Pediatr Infect Dis J [Internet]. 2018 Mar 1 [cited 2026 Mar 8];37(3):e58. Available from: https://pmc.ncbi.nlm.nih.gov/articles/PMC5807168/

31. Bar-Zeev N, Jere KC, Bennett A, Pollock L, Tate JE, Nakagomi O, et al. Population Impact and Effectiveness of Monovalent Rotavirus Vaccination in Urban Malawian Children 3 Years after Vaccine Introduction: Ecological and Case-Control Analyses. Clin Infect Dis [Internet]. 2016 May 1 [cited 2021 Jan 6];62(Suppl 2):S213–9. Available from: /pmc/articles/PMC4825885/?report=abstract

32. Beres LK, Tate JE, Njobvu L, Chibwe B, Rudd C, Guffey MB, et al. A Preliminary Assessment of Rotavirus Vaccine Effectiveness in Zambia. Clin Infect Dis [Internet]. 2016 May 1 [cited 2023 Feb 23];62 Suppl 2:S175–82. Available from: https://pubmed.ncbi.nlm.nih.gov/27059353/

33. Jere KC, Bar-Zeev N, Chande A, Bennett A, Pollock L, Sanchez-Lopez PF, et al. Vaccine Effectiveness against DS-1–Like Rotavirus Strains in Infants with Acute Gastroenteritis, Malawi, 2013–2015. Emerg Infect Dis [Internet]. 2019 [cited 2026 Mar 8];25(9):1734. Available from: https://pmc.ncbi.nlm.nih.gov/articles/PMC6711242/

34. Armah G, Pringle K, Enweronu-Laryea CC, Ansong D, Mwenda JM, Diamenu SK, et al. Impact and Effectiveness of Monovalent Rotavirus Vaccine Against Severe Rotavirus Diarrhea in Ghana. Clin Infect Dis [Internet]. 2016 May 1 [cited 2026 Mar 8];62 Suppl 2(Suppl 2):S200–7. Available from: https://pubmed.ncbi.nlm.nih.gov/27059357/

35. Gastañaduy PA, Steenhoff AP, Mokomane M, Esona MD, Bowen MD, Jibril H, et al. Effectiveness of Monovalent Rotavirus Vaccine after Programmatic Implementation in Botswana: A Multisite Prospective Case-Control Study. Clin Infect Dis. 2016;62(Suppl 2):S161–7.

36. Platts-Mills JA, Amour C, Gratz J, Nshama R, Walongo T, Mujaga B, et al. Impact of Rotavirus Vaccine Introduction and Postintroduction Etiology of Diarrhea Requiring Hospital Admission in Haydom, Tanzania, a Rural African Setting. Clin Infect Dis [Internet]. 2017 Oct 1 [cited 2026 Mar 8];65(7):1144–51. Available from: https://pubmed.ncbi.nlm.nih.gov/28575304/

37. Groome MJ, Page N, Cortese MM, Moyes J, Zar HJ, Kapongo CN, et al. Effectiveness of monovalent human rotavirus vaccine against admission to hospital for acute rotavirus diarrhoea in South African children: A case-control study. Lancet Infect Dis. 2014;14(11):1096–104.

38. Bar-Zeev N, Kapanda L, Tate JE, Jere KC, Iturriza-Gomara M, Nakagomi O, et al. Effectiveness of a monovalent rotavirus vaccine in infants in Malawi after programmatic roll-out: An observational and case-control study. Lancet Infect Dis [Internet]. 2015 Apr 1 [cited 2021 Jan 6];15(4):422–8. Available from: /pmc/articles/PMC4374102/?report=abstract

39. Abeid KA, Jani B, Cortese MM, Kamugisha C, Mwenda JM, Pandu AS, et al. Monovalent Rotavirus Vaccine Effectiveness and Impact on Rotavirus Hospitalizations in Zanzibar, Tanzania: Data From the First 3 Years After Introduction. J Infect Dis. 2017 Jan;215(2):183–91.

40. Bar-Zeev N, King C, Phiri T, Beard J, Mvula H, Crampin AC, et al. Impact of monovalent rotavirus vaccine on diarrhoea-associated post-neonatal infant mortality in rural communities in Malawi: a population-based birth cohort study. Lancet Glob Heal. 2018 Sep;6(9):e1036–44.

41. Enane LA, Gastañaduy PA, Goldfarb DM, Pernica JM, Mokomane M, Moorad B, et al. Impact of Rotavirus Vaccination on Hospitalizations and Deaths from Childhood Gastroenteritis in Botswana. Clin Infect Dis [Internet]. 2016 May 1 [cited 2021 Jan 6];62(Suppl 2):S168–74. Available from: /pmc/articles/PMC4825886/?report=abstract

42. Tsolenyanu E, Djadou KE, Fiawoo M, Akolly DAE, Mwenda JM, Leshem E, et al. Evidence of the impact of monovalent rotavirus vaccine on childhood acute gastroenteritis hospitalization in Togo. Vaccine [Internet]. 2018;36(47):7185–91. Available from: 10.1016/j.vaccine.2018.01.058

43. de Deus N, Chilaúle JJ, Cassocera M, Bambo M, Langa JS, Sitoe E, et al. Early impact of rotavirus vaccination in children less than five years of age in Mozambique. Vaccine [Internet]. 2018 Nov 12 [cited 2026 Mar 8];36(47):7205–9. Available from: https://pubmed.ncbi.nlm.nih.gov/29128381/

44. Rahajamanana VL, Raboba JL, Rakotozanany A, Razafindraibe NJ, Andriatahirintsoa EJPR, Razafindrakoto AC, et al. Impact of rotavirus vaccine on all-cause diarrhea and rotavirus hospitalizations in Madagascar. Vaccine [Internet]. 2017 Nov 12 [cited 2026 Mar 8];36(47):7198. Available from: https://pmc.ncbi.nlm.nih.gov/articles/PMC5867203/

45. Tsolenyanu E, Djadou KE, Fiawoo M, Akolly DAE, Mwenda JM, Leshem E, et al. Evidence of the impact of monovalent rotavirus vaccine on childhood acute gastroenteritis hospitalization in Togo. Vaccine [Internet]. 2018 Nov 12 [cited 2026 Mar 8];36(47):7185. Available from: https://pmc.ncbi.nlm.nih.gov/articles/PMC11973829/

46. Jani B, Hokororo A, Mchomvu J, Cortese MM, Kamugisha C, Mujuni D, et al. Detection of rotavirus before and after monovalent rotavirus vaccine introduction and vaccine effectiveness among children in mainland Tanzania. Vaccine [Internet]. 2018 Nov 12 [cited 2026 Mar 8];36(47):7149–56. Available from: https://pubmed.ncbi.nlm.nih.gov/29655631/

47. Mphahlele J, Abebe A, Getahun M, Sl M, Beyene B, Assefa E, et al. Impact of rotavirus vaccine introduction and genotypic. 2018;2011–6.

48. Sibomana H, Rugambwa C, Mwenda JM, Sayinzoga F, Iraguha G, Uwimana J, et al. Impact of routine rotavirus vaccination on all-cause and rotavirus hospitalizations during the first four years following vaccine introduction in Rwanda. Vaccine [Internet]. 2018;36(47):7135–41. Available from: 10.1016/j.vaccine.2018.01.072

49. Mpabalwani EM, Simwaka CJ, Mwenda JM, Mubanga CP, Monze M, Matapo B, et al. Impact of Rotavirus Vaccination on Diarrheal Hospitalizations in Children Aged <5 Years in Lusaka, Zambia. Clin Infect Dis [Internet]. 2016 May 1 [cited 2021 Jan 6];62(suppl 2):S183–7. Available from: https://academic.oup.com/cid/article-lookup/doi/10.1093/cid/civ1027

50. Enweronu-Laryea CC, Armah G, Sagoe KW, Ansong D, Addo-Yobo E, Diamenu SK, et al. Sustained impact of rotavirus vaccine introduction on rotavirus gastroenteritis hospitalizations in children <5 years of age, Ghana, 2009–2016. Vaccine [Internet]. 2018 Nov 12 [cited 2026 Mar 8];36(47):7131. Available from: https://pmc.ncbi.nlm.nih.gov/articles/PMC8848125/

51. Ahmed MLC brahim, Heukelbach J, Weddih A, Filali-Maltouf A, Sidatt M, Makhalla K, et al. Reduction of hospitalizations with diarrhea among children aged 0–5 years in Nouakchott, Mauritania, following the introduction of rotavirus vaccine. Vaccine [Internet]. 2019 Mar 7 [cited 2026 Mar 8];37(11):1407. Available from: https://pmc.ncbi.nlm.nih.gov/articles/PMC11726317/

52. Diop A, Thiongane A, Mwenda JM, Aliabadi N, Sonko MA, Diallo A, et al. Impact of rotavirus vaccine on acute gastroenteritis in children under 5 years in Senegal: Experience of sentinel site of the Albert Royer Children’s Hospital in Dakar. Vaccine [Internet]. 2018 Nov 12 [cited 2026 Mar 8];36(47):7192–7. Available from: https://pubmed.ncbi.nlm.nih.gov/29162319/

53. Mpabalwani EM, Simwaka JC, Mwenda JM, Matapo B, Parashar UD, Tate JE. Sustained impact of rotavirus vaccine on rotavirus hospitalisations in Lusaka, Zambia, 2009–2016. Vaccine [Internet]. 2018 Nov 12 [cited 2026 Mar 8];36(47):7165–9. Available from: https://pubmed.ncbi.nlm.nih.gov/29793891/

54. Maphalala G, Phungwayo N, Masona G, Lukhele N, Tsegaye G, Dube N, et al. Early impact of rotavirus vaccine in under 5 year old children hospitalized due to diarrhea, Swaziland. Vaccine [Internet]. 2018;36(47):7210–4. Available from: 10.1016/j.vaccine.2017.07.072

55. Bennett A, Pollock L, Jere KC, Pitzer VE, Parashar U, Tate JE, et al. Direct and possible indirect effects of vaccination on rotavirus hospitalisations among children in Malawi four years after programmatic introduction. Vaccine [Internet]. 2018 Nov 11 [cited 2024 Jul 9];36(47):7142. Available from: /pmc/articles/PMC6238204/

56. Mujuru HA, Burnett E, Nathoo KJ, Ticklay I, Gonah NA, Mukaratirwa A, et al. Monovalent Rotavirus Vaccine Effectiveness Against Rotavirus Hospitalizations Among Children in Zimbabwe. Clin Infect Dis [Internet]. 2019 Sep 27 [cited 2026 Mar 8];69(8):1339. Available from: https://pmc.ncbi.nlm.nih.gov/articles/PMC8851377/

57. Groome MJ, Zell ER, Solomon F, Nzenze S, Parashar UD, Izu A, et al. Temporal Association of Rotavirus Vaccine Introduction and Reduction in All-Cause Childhood Diarrheal Hospitalizations in South Africa. Clin Infect Dis [Internet]. 2016 May 1 [cited 2026 Mar 8];62(Suppl 2):S188. Available from: https://pmc.ncbi.nlm.nih.gov/articles/PMC11345717/

58. Wandera EA, Mohammad S, Bundi M, Nyangao J, Galata A, Kathiiko C, et al. Impact of rotavirus vaccination on rotavirus hospitalisation rates among a resource-limited rural population in Mbita, Western Kenya. Trop Med Int Health [Internet]. 2018 Apr 1 [cited 2026 Mar 8];23(4):425–32. Available from: https://pubmed.ncbi.nlm.nih.gov/29432666/

59. Madhi SA, Groome MJ, Zar HJ, Kapongo CN, Mulligan C, Nzenze S, et al. Effectiveness of pneumococcal conjugate vaccine against presumed bacterial pneumonia hospitalisation in HIV-uninfected South African children: a case-control study. Thorax [Internet]. 2015 Dec 1 [cited 2026 Mar 8];70(12):1149–55. Available from: https://pubmed.ncbi.nlm.nih.gov/26092924/

60. Izu A, Solomon F, Nzenze SA, Mudau A, Zell E, O’Brien KL, et al. Pneumococcal conjugate vaccines and hospitalization of children for pneumonia: a time-series analysis, South Africa, 2006–2014. Bull World Health Organ [Internet]. 2017 Sep 1 [cited 2026 Mar 8];95(9):618. Available from: https://pmc.ncbi.nlm.nih.gov/articles/PMC5578378/

61. Cohen C, Von Mollendorf C, De Gouveia L, Naidoo N, Meiring S, Quan V, et al. Effectiveness of 7-Valent Pneumococcal Conjugate Vaccine Against Invasive Pneumococcal Disease in HIV-Infected and –Uninfected Children in South Africa: A Matched Case-Control Study. Clin Infect Dis [Internet]. 2014 Sep 15 [cited 2026 Mar 8];59(6):808–18. Available from: 10.1093/cid/ciu431

62. Mackenzie GA, Hill PC, Jeffries DJ, Ndiaye M, Sahito SM, Hossain I, et al. Impact of the introduction of pneumococcal conjugate vaccination on invasive pneumococcal disease and pneumonia in The Gambia: 10 years of population-based surveillance. Lancet Infect Dis [Internet]. 2021 Sep 1 [cited 2026 Mar 8];21(9):1293–302. Available from: https://pubmed.ncbi.nlm.nih.gov/34280357/

63. Kambiré D, Soeters HM, Ouédraogo-Traoré R, Medah I, Sangaré L, Yaméogo I, et al. Early impact of 13-valent pneumococcal conjugate vaccine on pneumococcal meningitis-Burkina Faso, 2014-2015. J Infect. 2018 Mar;76(3):270–9.

64. Faye PM, Sonko MA, Diop A, Thiongane A, Ba ID, Spiller M, et al. Impact of 13-Valent Pneumococcal Conjugate Vaccine on Meningitis and Pneumonia Hospitalizations in Children aged <5 Years in Senegal, 2010-2016. Clin Infect Dis [Internet]. 2019 Sep 5 [cited 2026 Mar 8];69(Suppl 2):S66–71. Available from: https://pubmed.ncbi.nlm.nih.gov/31505625/

65. Cohen C, von Mollendorf C, de Gouveia L, Lengana S, Meiring S, Quan V, et al. Effectiveness of the 13-valent pneumococcal conjugate vaccine against invasive pneumococcal disease in South African children: a case-control study. Lancet Glob Heal [Internet]. 2017 Mar 1 [cited 2026 Mar 8];5(3):e359–69. Available from: https://pubmed.ncbi.nlm.nih.gov/28139443/

66. Dondo V, Mujuru H, Nathoo K, Jacha V, Tapfumanei O, Chirisa P, et al. Pneumococcal Conjugate Vaccine Impact on Meningitis and Pneumonia Among Children Aged <5 Years-Zimbabwe, 2010-2016. Clin Infect Dis [Internet]. 2019 Sep 5 [cited 2026 Mar 8];69(Suppl 2):S72–80. Available from: https://pubmed.ncbi.nlm.nih.gov/31505631/

67. Nhantumbo AA, Weldegebriel G, Katsande R, De Gouveia L, Comé CE, Cuco AZ, et al. Surveillance of impact of PCV-10 vaccine on pneumococcal meningitis in Mozambique, 2013 – 2015. PLoS One [Internet]. 2017 Jun 1 [cited 2026 Mar 8];12(6):e0177746. Available from: https://journals.plos.org/plosone/article?id=10.1371/journal.pone.0177746

68. Jenkins HE, Aylward RB, Gasasira A, Donnelly CA, Abanida EA, Koleosho-Adelekan T, et al. Effectiveness of immunization against paralytic poliomyelitis in Nigeria. N Engl J Med [Internet]. 2008 Oct 16 [cited 2026 Mar 8];359(16):1666–74. Available from: https://pubmed.ncbi.nlm.nih.gov/18923171/

69. Mahamud A, Kamadjeu R, Webeck J, Mbaeyi C, Baranyikwa MT, Birungi J, et al. Effectiveness of oral polio vaccination against paralytic poliomyelitis: a matched case-control study in Somalia. J Infect Dis. 2014 Nov;210 Suppl:S187–93.

70. Chippaux J-P, Diallo A, Marra A, Etard J-F. Impact of previous immunisation on the incidence of meningococcal disease during an outbreak in a Sahelian area of Senegal. Vaccine. 2007 Feb;25(10):1712–8.

71. Kristiansen PA, Diomandé F, Ba AK, Sanou I, Ouédraogo AS, Ouédraogo R, et al. Impact of the serogroup A meningococcal conjugate vaccine, MenAfriVac, on carriage and herd immunity. Clin Infect Dis [Internet]. 2013 Feb [cited 2026 Mar 8];56(3):354–63. Available from: https://pubmed.ncbi.nlm.nih.gov/23087396/

72. Gamougam K, Daugla DM, Toralta J, Ngadoua C, Fermon F, Page A-L, et al. Continuing effectiveness of serogroup A meningococcal conjugate vaccine, Chad, 2013. Emerg Infect Dis. 2015 Jan;21(1):115–8.

73. Sugerman DE, Fall A, Guigui M-T, N’dolie M, Balogun T, Wurie A, et al. Preplanned national measles vaccination campaign at the beginning of a measles outbreak--Sierra Leone, 2009-2010. J Infect Dis. 2011 Jul;204 Suppl:S260–9.

74. Kidd S, Ouedraogo B, Kambire C, Kambou JL, McLean H, Kutty PK, et al. Measles outbreak in Burkina Faso, 2009: a case-control study to determine risk factors and estimate vaccine effectiveness. Vaccine. 2012 Jul;30(33):5000–8.

75. Majwala RK, Nakiire L, Kadobera D, Ario AR, Kusiima J, Atuhairwe JA, et al. Measles outbreak propagated by children congregating at PHSMs collection points in Mayuge District, eastern Uganda, July – October, 2016. BMC Infect Dis. 2018 Aug;18(1):412.

76. Mupere E, Karamagi C, Zirembuzi G, Grabowsky M, de Swart RL, Nanyunja M, et al. Measles vaccination effectiveness among children under 5 years of age in Kampala, Uganda. Vaccine. 2006 May;24(19):4111–5.

77. Goodson JL, Wiesen E, Perry RT, Mach O, Kitambi M, Kibona M, et al. Impact of measles outbreak response vaccination campaign in Dar es Salaam, Tanzania. Vaccine. 2009 Sep;27(42):5870–4.

78. Doshi RH, Shidi C, Mulumba A, Eckhoff P, Nguyen C, Hoff NA, et al. The effect of immunization on measles incidence in the Democratic Republic of Congo: Results from a model of surveillance data. Vaccine [Internet]. 2015 Nov 27 [cited 2026 Mar 8];33(48):6786–92. Available from: https://pubmed.ncbi.nlm.nih.gov/26476363/

79. Minetti A, Kagoli M, Katsulukuta A, Huerga H, Featherstone A, Chiotcha H, et al. Lessons and challenges for measles control from unexpected large outbreak, Malawi. Emerg Infect Dis. 2013;19(2):202–9.

80. Doshi RH, Mukadi P, Shidi C, Mulumba A, Hoff NA, Gerber S, et al. Field evaluation of measles vaccine effectiveness among children in the Democratic Republic of Congo. Vaccine [Internet]. 2015 Jun 26 [cited 2026 Mar 8];33(29):3407–14. Available from: https://www.sciencedirect.com/science/article/abs/pii/S0264410X15005435

81. Katz MA, Lebo E, Emukule GO, Otieno N, Caselton DL, Bigogo G, et al. Uptake and Effectiveness of a Trivalent Inactivated Influenza Vaccine in Children in Urban and Rural Kenya, 2010 to 2012. Pediatr Infect Dis J [Internet]. 2016 Mar 1 [cited 2026 Mar 8];35(3):322–9. Available from: https://pubmed.ncbi.nlm.nih.gov/26658627/

82. Ntshoe GM, McAnerney JM, Tempia S, Blumberg L, Moyes J, Buys A, et al. Influenza Epidemiology and Vaccine Effectiveness among Patients with Influenza-Like Illness, Viral Watch Sentinel Sites, South Africa, 2005–2009. PLoS One [Internet]. 2014 Apr 15 [cited 2026 Mar 8];9(4):e94681. Available from: https://journals.plos.org/plosone/article?id=10.1371/journal.pone.0094681

83. Hansen OB, Rodrigues A, Martins C, Rieckmann A, Benn CS, Aaby P, et al. Impact of H1N1 Influenza Vaccination on Child Morbidity in Guinea-Bissau. J Trop Pediatr [Internet]. 2019 Oct 1 [cited 2026 Mar 8];65(5):446–56. Available from: 10.1093/tropej/fmy075

84. McAnerney JM, Walaza S, Tempia S, Blumberg L, Treurnicht FK, Madhi SA, et al. Estimating vaccine effectiveness in preventing laboratory-confirmed influenza in outpatient settings in South Africa, 2015. Influenza Other Respi Viruses [Internet]. 2017 Mar 1 [cited 2026 Mar 8];11(2):177–81. Available from: https://pubmed.ncbi.nlm.nih.gov/27865064/

85. McAnerney JM, Walaza S, Cohen AL, Tempia S, Buys A, Venter M, et al. Effectiveness and knowledge, attitudes and practices of seasonal influenza vaccine in primary healthcare settings in South Africa, 2010-2013. Influenza Other Respi Viruses. 2015 May;9(3):143–50.

86. Lee EHJ, Lewis RF, Makumbi I, Kekitiinwa A, Ediamu TD, Bazibu M, et al. Haemophilus influenzae type b conjugate vaccine is highly effective in the Ugandan routine immunization program: a case-control study. Trop Med Int Health [Internet]. 2008 Apr [cited 2026 Mar 8];13(4):495–502. Available from: https://pubmed.ncbi.nlm.nih.gov/18312475/

87. Madhi SA, Petersen K, Khoosal M, Huebner RE, Mbelle N, Mothupi R, et al. Reduced effectiveness of Haemophilus influenzae type b conjugate vaccine in children with a high prevalence of human immunodeficiency virus type 1 infection. Pediatr Infect Dis J [Internet]. 2002 [cited 2026 Mar 8];21(4):315–21. Available from: https://pubmed.ncbi.nlm.nih.gov/12075763/

88. Fleming JA, Dieye Y, Ba O, Mutombo Wa Mutombo B, Diallo N, Faye PC, et al. Effectiveness of haemophilus influenzae type B conjugate vaccine for prevention of meningitis in Senegal. Pediatr Infect Dis J [Internet]. 2011 [cited 2026 Mar 8];30(5):430–2. Available from: https://pubmed.ncbi.nlm.nih.gov/21099444/

89. Cowgill KD, Ndiritu M, Nyiro J, Slack MPE, Chiphatsi S, Ismail A, et al. Effectiveness of Haemophilus influenzae type b conjugate vaccine introduction into routine childhood immunization in Kenya. JAMA [Internet]. 2006 Aug 9 [cited 2026 Mar 8];296(6):671. Available from: https://pmc.ncbi.nlm.nih.gov/articles/PMC1592684/

90. Lucas MES, Deen JL, von Seidlein L, Wang X-Y, Ampuero J, Puri M, et al. Effectiveness of mass oral cholera vaccination in Beira, Mozambique. N Engl J Med [Internet]. 2005 Feb 24 [cited 2026 Mar 8];352(8):757–67. Available from: https://pubmed.ncbi.nlm.nih.gov/15728808/

91. Luquero FJ, Grout L, Ciglenecki I, Sakoba K, Traore B, Heile M, et al. Use of Vibrio cholerae vaccine in an outbreak in Guinea. N Engl J Med [Internet]. 2014 May 29 [cited 2026 Mar 8];370(22):2111–20. Available from: https://pubmed.ncbi.nlm.nih.gov/24869721/

92. Tate JE, Ngabo F, Donnen P, Gatera M, Uwimana J, Rugambwa C, et al. Effectiveness of Pentavalent Rotavirus Vaccine Under Conditions of Routine Use in Rwanda. Clin Infect Dis [Internet]. 2016 May 1 [cited 2026 Mar 8];62 Suppl 2(Suppl 2):S208–12. Available from: https://pubmed.ncbi.nlm.nih.gov/27059358/

93. Bar-zeev N, Jere KC, Bennett A, Pollock L, Tate JE, Nakagomi O, et al. Population Impact and Effectiveness of Monovalent Rotavirus Vaccination in Urban Malawian Children 3 Years After Vaccine IntroductionlJ: Ecological and Case-Control Analyses. 2016;62(Suppl 2):213–9.

94. Armah G, Pringle K, Enweronu-Laryea CC, Ansong D, Mwenda JM, Diamenu SK, et al. Impact and Effectiveness of Monovalent Rotavirus Vaccine Against Severe Rotavirus Diarrhea in Ghana. Clin Infect Dis [Internet]. 2016 May 1 [cited 2023 Feb 21];62 Suppl 2(Suppl 2):S200–7. Available from: https://pubmed.ncbi.nlm.nih.gov/27059357/

95. Groome MJ, Page N, Cortese MM, Moyes J, Zar HJ, Kapongo CN, et al. Effectiveness of monovalent human rotavirus vaccine against admission to hospital for acute rotavirus diarrhoea in South African children: A case-control study. Lancet Infect Dis [Internet]. 2014 Nov 1 [cited 2026 Mar 8];14(11):1096–104. Available from: https://pubmed.ncbi.nlm.nih.gov/25303843/

96. Mujuru HA, Burnett E, Nathoo KJ, Ticklay I, Gonah NA, Mukaratirwa A, et al. Monovalent Rotavirus Vaccine Effectiveness Against Rotavirus Hospitalizations Among Children in Zimbabwe. Clin Infect Dis an Off Publ Infect Dis Soc Am. 2019 Sep;69(8):1339–44.

97. Sibomana H, Rugambwa C, Mwenda JM, Sayinzoga F, Iraguha G, Uwimana J, et al. Impact of routine rotavirus vaccination on all-cause and rotavirus hospitalizations during the first four years following vaccine introduction in Rwanda. Vaccine [Internet]. 2018 Nov 12 [cited 2026 Mar 8];36(47):7135. Available from: https://pmc.ncbi.nlm.nih.gov/articles/PMC11973826/

98. Enweronu-Laryea CC, Armah G, Sagoe KW, Ansong D, Addo-Yobo E, Diamenu SK, et al. Sustained impact of rotavirus vaccine introduction on rotavirus gastroenteritis hospitalizations in children <5 years of age, Ghana, 2009–2016. Vaccine [Internet]. 2018 Nov 12 [cited 2021 Jan 6];36(47):7131–4. Available from: https://pubmed.ncbi.nlm.nih.gov/29752020/

99. Ahmed M-LC-B, Heukelbach J, Weddih A, Filali-Maltouf A, Sidatt M, Makhalla K, et al. Reduction of hospitalizations with diarrhea among children aged 0-5 years in Nouakchott, Mauritania, following the introduction of rotavirus vaccine. Vaccine. 2019 Mar;37(11):1407–11.

100. Mpabalwani EM, Simwaka JC, Mwenda JM, Matapo B, Parashar UD, Tate JE. Sustained impact of rotavirus vaccine on rotavirus hospitalisations in Lusaka, Zambia, 2009-2016. Vaccine [Internet]. 2018 Nov 12 [cited 2023 Feb 22];36(47):7165–9. Available from: https://pubmed.ncbi.nlm.nih.gov/29793891/

101. Bennett A, Pollock L, Jere KC, Pitzer VE, Parashar U, Tate JE, et al. Direct and possible indirect effects of vaccination on rotavirus hospitalisations among children in Malawi four years after programmatic introduction. Vaccine. 2018 Nov;36(47):7142–8.

102. Wandera EA, Mohammad S, Bundi M, Nyangao J, Galata A, Kathiiko C, et al. Impact of rotavirus vaccination on rotavirus hospitalisation rates among a resource-limited rural population in Mbita, Western Kenya. Trop Med Int Health [Internet]. 2018 Apr 1 [cited 2023 Feb 22];23(4):425–32. Available from: https://pubmed.ncbi.nlm.nih.gov/29432666/

103. Enane LA, Gastañaduy PA, Goldfarb DM, Pernica JM, Mokomane M, Moorad B, et al. Impact of Rotavirus Vaccination on Hospitalizations and Deaths From Childhood Gastroenteritis in Botswana. Clin Infect Dis [Internet]. 2016 May 1 [cited 2026 Mar 8];62(suppl_2):S168–74. Available from: 10.1093/cid/civ1210

104. Rahajamanana VL, Raboba JL, Rakotozanany A, Razafindraibe NJ, Andriatahirintsoa EJPR, Razafindrakoto AC, et al. Impact of rotavirus vaccine on all-cause diarrhea and rotavirus hospitalizations in Madagascar. Vaccine. 2018 Nov;36(47):7198–204.

105. Tsolenyanu E, Mwenda JM, Dagnra A, Leshem E, Godonou M, Nassoury I, et al. Early Evidence of Impact of Monovalent Rotavirus Vaccine in Togo. Clin Infect Dis [Internet]. 2016 May 1 [cited 2026 Mar 8];62 Suppl 2:S196–9. Available from: https://pubmed.ncbi.nlm.nih.gov/27059356/

106. Bar-Zeev N, Jere KC, Bennett A, Pollock L, Tate JE, Nakagomi O, et al. Population Impact and Effectiveness of Monovalent Rotavirus Vaccination in Urban Malawian Children 3 Years After Vaccine Introduction: Ecological and Case-Control Analyses. Clin Infect Dis [Internet]. 2016 May 1 [cited 2026 Mar 8];62 Suppl 2(Suppl 2):S213–9. Available from: https://pubmed.ncbi.nlm.nih.gov/27059359/

107. Soeters HM, Kambiré D, Sawadogo G, Ouédraogo-Traoré R, Bicaba B, Medah I, et al. Impact of 13-Valent Pneumococcal Conjugate Vaccine on Pneumococcal Meningitis, Burkina Faso, 2016-2017. J Infect Dis [Internet]. 2019 Oct 31 [cited 2026 Mar 8];220(220 Suppl 4):S253–62. Available from: https://pubmed.ncbi.nlm.nih.gov/31671444/

108. Faye PM, Sonko MA, Diop A, Thiongane A, Ba ID, Spiller M, et al. Impact of 13-Valent Pneumococcal Conjugate Vaccine on Meningitis and Pneumonia Hospitalizations in Children aged <5 Years in Senegal, 2010-2016. Clin Infect Dis [Internet]. 2019 Sep 5 [cited 2020 Dec 18];69(Suppl 2):S66–71. Available from: /pmc/articles/PMC6756160/?report=abstract

109. Dondo V, Mujuru H, Nathoo K, Jacha V, Tapfumanei O, Chirisa P, et al. Pneumococcal Conjugate Vaccine Impact on Meningitis and Pneumonia Among Children Aged <5 Years-Zimbabwe, 2010-2016. Clin Infect Dis an Off Publ Infect Dis Soc Am. 2019 Sep;69(Suppl 2):S72–80.

110. Mupere E, Karamagi C, Zirembuzi G, Grabowsky M, de Swart RL, Nanyunja M, et al. Measles vaccination effectiveness among children under 5 years of age in Kampala, Uganda. Vaccine [Internet]. 2006 May 8 [cited 2026 Mar 8];24(19):4111–5. Available from: https://pubmed.ncbi.nlm.nih.gov/16554111/

111. Sugerman DE, Fall A, Guigui MT, N’dolie M, Balogun T, Wurie A, et al. Preplanned national measles vaccination campaign at the beginning of a measles outbreak--Sierra Leone, 2009-2010. J Infect Dis [Internet]. 2011 Jul 1 [cited 2026 Mar 8];204 Suppl 1(SUPPL. 1). Available from: https://pubmed.ncbi.nlm.nih.gov/21666171/

112. Ntshoe GM, McAnerney JM, Tempia S, Blumberg L, Moyes J, Buys A, et al. Influenza epidemiology and vaccine effectiveness among patients with influenza-like illness, viral watch sentinel sites, South Africa, 2005-2009. PLoS One. 2014;9(4):e94681.

113. McAnerney JM, Walaza S, Cohen AL, Tempia S, Buys A, Venter M, et al. Effectiveness and knowledge, attitudes and practices of seasonal influenza vaccine in primary healthcare settings in South Africa, 2010–2013. Influenza Other Respi Viruses [Internet]. 2015 May 1 [cited 2026 Mar 8];9(3):143. Available from: https://pmc.ncbi.nlm.nih.gov/articles/PMC4415698/

114. Chippaux JP, Diallo A, Marra A, Étard JF. Impact of previous immunisation on the incidence of meningococcal disease during an outbreak in a Sahelian area of Senegal. Vaccine [Internet]. 2007 Feb 26 [cited 2026 Mar 8];25(10):1712–8. Available from: https://pubmed.ncbi.nlm.nih.gov/17240492/

115. Gamougam K, Daugla DM, Toralta J, Ngadoua C, Fermon F, Page AL, et al. Continuing effectiveness of serogroup A meningococcal conjugate vaccine, Chad, 2013. Emerg Infect Dis [Internet]. 2015 [cited 2026 Mar 8];21(1):115–8. Available from: https://pubmed.ncbi.nlm.nih.gov/25536336/

116. Mahamud A, Kamadjeu R, Webeck J, Mbaeyi C, Baranyikwa MT, Birungi J, et al. Effectiveness of oral polio vaccination against paralytic poliomyelitis: a matched case-control study in Somalia. J Infect Dis [Internet]. 2014 Nov 1 [cited 2026 Mar 8];210 Suppl 1:S187–93. Available from: https://pubmed.ncbi.nlm.nih.gov/25316835/

117. Ndeketa L, Haine V, Debois M, Asante KP, Agyapong PD, Kaali S, et al. Effectiveness of the RTS,S/AS01E malaria vaccine in a real-world setting over 1 year of follow-up after the three-dose primary schedule: an interim analysis of a phase 4 study in Ghana, Kenya, and Malawi. Lancet Glob Heal [Internet]. 2025 Jan 1 [cited 2026 Jan 7];14(1):e61–9. Available from: https://www.thelancet.com/action/showFullText?pii=S2214109X25004152

118. Bar-Zeev N, Jere KC, Bennett A, Pollock L, Tate JE, Nakagomi O, et al. Population Impact and Effectiveness of Monovalent Rotavirus Vaccination in Urban Malawian Children 3 Years After Vaccine Introduction: Ecological and Case-Control Analyses. Clin Infect Dis [Internet]. 2016 May 1 [cited 2023 Feb 23];62 Suppl 2(Suppl 2):S213–9. Available from: https://pubmed.ncbi.nlm.nih.gov/27059359/

119. Ahmed ST, Haider SS, Hanif S, Anwar HB, Mehjabeen S, Closser S, et al. A scoping review on integrated health campaigns for immunization in low– and middle-income countries. Health Policy Plan [Internet]. 2023 Nov 28 [cited 2026 Jan 14];38(10):1198–224. Available from: 10.1093/heapol/czad082

120. Grimes DA, Schulz KF. Cohort studies: Marching towards outcomes. Lancet [Internet]. 2002 Jan 26 [cited 2026 Jan 14];359(9303):341–5. Available from: https://www.thelancet.com/action/showFullText?pii=S0140673602075001

121. Izu A, Solomon F, Nzenze SA, Mudau A, Zell E, O’Brien KL, et al. Pneumococcal conjugate vaccines and hospitalization of children for pneumonia: a time-series analysis, South Africa, 2006–2014. Bull World Health Organ [Internet]. 2017 Sep 1 [cited 2025 Jul 2];95(9):618. Available from: https://pmc.ncbi.nlm.nih.gov/articles/PMC5578378/

122. Asante KP, Mathanga DP, Milligan P, Akech S, Oduro A, Mwapasa V, et al. Feasibility, safety, and impact of the RTS,S/AS01E malaria vaccine when implemented through national immunisation programmes: evaluation of cluster-randomised introduction of the vaccine in Ghana, Kenya, and Malawi. Lancet [Internet]. 2024 Apr 27 [cited 2026 Jan 14];403(10437):1660–70. Available from: https://www.sciencedirect.com/science/article/pii/S0140673624000047

123. Ozawa S, Clark S, Portnoy A, Grewal S, Brenzel L, Walker DG. Return On Investment From Childhood Immunization In Low– And Middle-Income Countries, 2011–20. 101377/hlthaff20151086 [Internet]. 2017 Aug 2 [cited 2026 Jan 15];35(2):199–207. Available from: /doi/pdf/10.1377/hlthaff.2015.1086

124. Sallam M, Sallam M. COVID-19 Vaccine Hesitancy Worldwide: A Concise Systematic Review of Vaccine Acceptance Rates. Vaccines 2021, Vol 9, [Internet]. 2021 Feb 16 [cited 2026 Jan 15];9(2):1–15. Available from: https://www.mdpi.com/2076-393X/9/2/160

