## Supplementaty table for "Methods of adjustment for non-vaccine interventions in post-licensure vaccine studies in children in sub–Saharan Africa: a systematic review"

**Supplementary Table S1: Summary of vaccine effectiveness and impact patterns across included studies stratified by pathogen and study design.**

| **Pathogen** | **Studies (n)** | **Design (n)** | **Vaccine** | **Primary End Points** | **Key Impact Findings** | **Notable Patterns** |
| --- | --- | --- | --- | --- | --- | --- |
| Rotavirus | 31 | Ecological (15)  Case control (12)  Time series (3)  Cohort (1) | RV1, RV5 | Hospitalizations for rotavirus gastroenteritis | 30–76% population impact in hospitalisations post-vaccination | Lower VE in high-HIV/malnourished populations  RV1 n=28, RV5 n=3 |
| Streptococcus pneumoniae | 9 | Case control (4)  Ecological (2)  Time series (3) | PCV7/10/13 | Prevention of invasive pneumococcal disease (IPD) and pneumonia hospitalizations | 52-80% reduction in invasive pneumococcal disease incidence. 27-39% decrease in pneumonia hospitalizations. 80-91% effectiveness against vaccine-type IPD in HIV-negative children | Higher VE against meningitis than pneumonia Lower effectiveness in HIV positive children (57% vs 90%) PCV13 shows broader protection than PCV7/10 |
| Measles | 8 | Ecological (4)  Case control (3)  Cohort (1) | Measles containing vaccine | Measles case prevention and outbreak control | 74-99% reduction in measles cases among vaccinated children 67-80% decrease in outbreak incidence following vaccination campaigns | Lower effectiveness in outbreak settings (74-75%) 2-dose regimens show consistently higher protection (84-99%) than single dose Effectiveness varies by study design (case-control vs ecological) |
| Influenza A/B (seasonal) | 5 | Case control (4)  Ecological (1) | Inactivated/ Tetravalent | Lab-confirmed outpatient cases | Season dependent effectiveness, weakest protection during mismatched seasons (negative VE) | Lower VE in high-HIV settings |
| Haemophilus influenzae type B | 4 | Case control (2)  Cohort (1)  Ecological (1) | Hib conjugate vaccine | Prevention of Hib meningitis & invasive disease | 87-99% reduction in Hib meningitis cases 98.8% protection against Hib meningitis deaths 88-96% effectiveness in routine immunisation programs 96.5% (74.4–99.5%) effectiveness in HIV positive children 43.9% (-76.1–82.1%) effectiveness in HIV positive children 5.9x higher baseline Hib risk in HIV positive infants | Lower effectiveness in HIV positive children  Near-elimination of Hib in high-coverage areas Sustained protection with 2-3 doses |
| Neisseria meningitidis (Serogroup A) | 3 | Ecological (1)  Times series (1) | MenAfriVac (PsA-tt conjugate) & Polysaccharide vaccine | Carriage reduction & meningitis prevention | 52-90% reduction in meningitis cases 67% decrease in carriage prevalence Herd protection observed within 1 year Polysaccharide vaccine has short acting protection in children and insufficient immunity in infants | Conjugate vaccine (MenAfriVac) has stronger protection than polysaccharide Protection lasts ≥3 years post-vaccination No serogroup replacement observed |
| Vibrio cholerae | 2 | Case control (2) | Oral cholera vaccine | Outbreak case reduction | 78-87% protection during outbreaks 2-dose effectiveness: 86.6% (56.7-95.8%) Single dose: 78% (39-92%) | High effectiveness in reactive campaigns Minimal protection for <12 month-olds Works best when combined with WASH interventions |
| Poliovirus (Types 1 & 3) | 2 | Case control (2) | OPV (monovalent & trivalent) | Paralytic polio prevention | mOPV1 showed 4x higher efficacy than tOPV against type 1 Minimal protection against type 3 with tOPV | Monovalent far outperforms trivalent vaccine Dose-dependent protection observed Study conducted during active wild polio transmission |
